# COVID-19 Vaccination in Pregnancy: The Impact of Multimorbidity and Smoking Status on Vaccine Hesitancy, a Cohort Study of 25,111 Women in Wales, UK

**DOI:** 10.1101/2022.12.12.22283200

**Authors:** M Mhereeg, H Jones, J Kennedy, M Seaborne, M Parker, N Kennedy, A Akbari, L Zuccolo, A Azcoaga-Lorenzo, A Davies, K Nirantharakumar, S Brophy

## Abstract

**Background:** Multimorbidity and pregnancy are two risk factors for more severe outcomes after a SARS-CoV-2 infection, thus vaccination uptake is important for pregnant women living with multimorbidity. This study aimed to examine the impact of multimorbidity, smoking status, and demographics (age, ethnic group, area of deprivation) on vaccine hesitancy among pregnant women in Wales using electronic health records (EHR) linkage.

**Methods:** This cohort study utilised routinely collected, individual-level, anonymised population-scale linked data within the Secure Anonymised Information Linkage (SAIL) Databank. Pregnant women were identified from 13^th^ April 2021 to 31^st^ December 2021. Survival analysis was utilised to examine and compare the length of time to vaccination uptake in pregnancy by multimorbidity and smoking status, as well as depression, diabetes, asthma, and cardiovascular conditions independently. Variation in uptake by; multimorbidity, smoking status, and demographics was examined jointly and separately for the independent conditions using hazard ratios (HR) from the Cox regression model. A bootstrapping internal validation was conducted to assess the performance of the models.

**Results:** Within the population cohort, 8,203 (32.7%) received at least one dose of the COVID-19 vaccine during pregnancy, with 8,572 (34.1%) remaining unvaccinated throughout the follow-up period, and 8,336 (33.2%) receiving the vaccine postpartum. Women aged 30 years or older were more likely to have the vaccine in pregnancy. Those who had depression were slightly but significantly more likely to have the vaccine compared to those without depression (HR = 1.08, 95% CI 1.03 to 1.14, p = 0.02). Women living with multimorbidity (> 1 health condition) were 1.12 times more likely to have the vaccine compared to those living without multimorbidity (HR = 1.12, 95% CI 1.04 to 1.19, p = 0.001). Vaccine uptakes were significantly lower among both current smokers and former smokers compared to never smokers (HR = 0.87, 95% CI 0.81 to 0.94, p < 0.001 and HR = 0.92, 95% CI 0.85 to 0.98, p = 0.015 respectively). Uptake was also lower among those living in the most deprived areas compared to those living in the most affluent areas (HR = 0.89, 95% CI 0.83 to 0.96, p = 0.002). The validated model had similar performance and revealed that multimorbidity, smoking status, age, and deprivation level together have a significant impact on vaccine hesitancy (p < 0.05 for all).

**Conclusion:** Younger women, living without multimorbidity (zero or only one health condition), current and former smokers, and those living in the more deprived areas are less likely to have the vaccine, thus, a targeted approach to vaccinations may be required for these groups. Women living with multimorbidity are slightly but significantly less likely to be hesitant about COVID-19 vaccination when pregnant.

## Background

COVID-19 vaccination is recognised as a successful public health measure [1], and national vaccine strategies were implemented during the COVID-19 pandemic. However, an increasing number of the general population perceive vaccinations as dangerous and not needed [1]. Vaccine hesitancy may be more prevalent in different populations [2]. For example, vaccine hesitancy may be more common in pregnant women as they have additional concerns [3]. During the COVID-19 pandemic, the limited data and changes in advice and recommendations regarding vaccination in pregnancy inevitably led to some hesitancy towards being vaccinated among pregnant women [4]. Despite increasing evidence that COVID-19 vaccines are safe and effective for pregnant women, vaccine hesitancy remains high [5].

Research is limited on population-level COVID-19 vaccine uptake in pregnancy in the UK. However, in Scotland, a national, prospective cohort study identifying ongoing pregnancies through extensive electronic health records (EHR) linkages showed vaccination rates in pregnant women to be considerably lower than in the general population; 32.3% in pregnant women compared to 77.4% in all women [6]. In England, 22.7% of women giving birth in August 2021 had received at least one dose of vaccine. This increased to 32.3% of women who gave birth in September and rose again to 53.7% in December 2021 [7]. In Wales, a mixed methods study utilising routinely collected linked data from the Secure Anonymised Information Linkage (SAIL) Databank and the Born In Wales Birth Cohort found that only 1 in 3 pregnant women would have the COVID-19 vaccine during pregnancy, even though 2 in 3 reported they would have the vaccination [5]. Despite the overall increase in coverage, the uptake remains lower amongst pregnant women compared to the general population of the same age group [7].

Various factors may influence vaccine acceptance or refusal. Multimorbidity is defined as the co-occurrence of two or more long-term health conditions, which can include defined physical and mental health conditions such as diabetes, depression, asthma, and cardiovascular diseases [8]. Long-term health conditions are those that generally last a year or longer and have a significant impact on a person’s life [9]. In the United Kingdom (UK), the COVID-19 pandemic has affected expectant mothers’ mental health by increasing the prevalence of depression by 47% [10]. Research has found that having pre-existing illnesses or multimorbidity is associated with a lower rate of vaccine refusal [11,12].

Health related behaviours may influence vaccine acceptance, such as smoking habits. Smoking status includes the categories of current smoker, former smoker, never smoked, or smoking status unknown. The differences in negative attitudes towards vaccines in general and intentions to vaccinate against COVID-19, specifically by smoking status, has been examined in a sample of adults in the UK [13]. Relative to never smoked and former smokers, current smokers reported significantly higher mistrust of vaccines and any benefits, were more worried about negative future effects, and had a stronger preference for natural immunity. With a high number of smokers belonging to socially clustered and disadvantaged socioeconomic groups, lower vaccine uptake in this group could also heighten health inequalities [13].

The determinants of vaccine hesitancy have been explored previously, aiming to create targeted interventions for the most at-risk groups. From 1,203 young adults aged 18-25 in the United States, vaccine hesitancy was significantly higher among young adults who were current smokers (including electronic cigarettes) when compared to non-current smokers (36% vs. 22%) [14]. Moreover, multivariable regression analysis demonstrated that current smokers, compared to non-current smokers, had more than two times the odds of reporting COVID-19 vaccine hesitancy [14]. In a similar study, the prevalence of COVID-19 vaccine hesitancy in the UK was analysed, which reported again that vaccine hesitancy was higher in younger age groups (26.5% in 16–24-year-olds compared to 4.5% in ages 75+) [15]. Vaccine acceptance also varied between ethnic groups. Vaccine hesitancy was higher in Black (71.8%) and Pakistani or Bangladeshi (42.3%) ethnic groups and lowest in the White British or Irish group (15.6%) [15].

The Office for National Statistics (ONS) investigated hesitancy towards the COVID-19 vaccine based on the Opinions and Lifestyle Survey (OPN) covering the period from 23^rd^ June to 18^th^ July 2021. It was reported that adults living in the most deprived areas of England (based on the Index of Multiple Deprivation) were more likely to report vaccine hesitancy (8%) than adults living in the least deprived areas (2%) [16].

The current study aims to examine the impact of multimorbidity, smoking status, and demographics (age, ethnic group, area of deprivation) on vaccine hesitancy among pregnant women in Wales using electronic health records (EHR) linkage. Identifying groups with higher vaccine hesitancy is critical to develop targeted interventions to enhance vaccine uptake rates.

## Methods

### Study design and setting

A cohort study utilising routinely collected individual-level, anonymised population-scale linked data within the Secure Anonymised Information Linkage (SAIL) Databank. Data sources include general practitioners (GP), hospital admissions, national community child health, maternal indicators, and vaccination data sources. All women recorded as being pregnant on or after 13^th^ April 2021, aged 18 years or older, and eligible for COVID-19 vaccination were identified. They were linked to the COVID-19 vaccination data for dates up to and including 31^st^ December 2021.

### Data sources and linkage

Analysis was undertaken using anonymised population-scale, individual-level linked routinely collected national-scale data available in the SAIL Databank [17,18], which anonymously links a wide range of person-based data employing a unique personal identifier. The linkage includes primary care data (from Wales Longitudinal General Practice (WLGP)) linked with secondary care data (from inpatient hospital admissions, inpatient from Patient Episode Database for Wales (PEDW) and outpatient from Outpatient Database for Wales (OPDW)), pregnancy and maternity related data (from the National Community Child Health (NCCH) and Maternal Indicators (MIDS)) and vaccination data (from the COVID-19 Vaccination Dataset (CVVD)). The primary care data utilises Read codes, which are predominantly 5-digit codes that relate to diagnosis, medication, and process of care codes. The secondary care data uses International Classification of Diseases version 10 (ICD-10) codes for diagnosis and OPCS Classification of Interventions and Procedures version 4 (OPCS-4) surgical interventions. The NCCH comprises information pertaining to birth registration, monitoring of child health examinations, and immunisations. The MIDS data contains data relating to the woman at initial assessment and to the mother and baby (or babies) for all births. In addition to these data sources, the Welsh Demographic Service Dataset (WDSD) was linked to extract Lower-layer Super Output Area (LSOA) version 2011 information associated with area-level deprivation. In particular, the Welsh Index for Multiple Deprivation (WIMD) version 2019 was employed as a proxy to assess socioeconomic status. These records were linked at the individual level for all women known to be pregnant in Wales between 13^th^ April 2021 and 31^st^ December 2021. Linkage quality has been assessed and reported as 99.9% for WLGP records and 99.3% for PEDW records [19]. All linkage was at the individual level.

### Study population and key dates

Pregnant women were identified as any woman who had pregnancy codes in the WLGP data, in hospital admissions (PEDW) for pregnancy, or mothers in the NCCH or MIDS data with the baby birth date (Pregnancy end date) and gestational age at birth available. The baby’s birth date and gestational age enabled the start date of pregnancy to be determined for those who gave birth during the study period. Data collected included vaccination data, maternal age, ethnic group, WIMD 2019, smoking status, depression, diabetes, asthma, and cardiovascular disease. The WIMD 2019 is an official measure for the relative deprivation of areas of Wales. It combines eight separate domains of deprivation, each compiled from a range of different indicators (income, employment, health, education, access to services, housing, community safety, and physical environment) into a single score and is widely utilised to measure deprivation in Wales. Ethnic groups are categorised in SAIL into White, Asian, Mixed, Black, and Other. Smoking status is categorised in SAIL into Current Smoker, Former Smoker, Never Smoker, and Unknown [22]. In cases where women had multiple recorded statuses, the most recent status during or prior to pregnancy was selected.

The study start date of 13^th^ April 2021 was selected because phase 2 of the vaccination program, which aimed to provide vaccinations to individuals aged 40 to 49, 30 to 39, and 18 to 29 years, commenced on this date. The inclusion criteria were currently pregnant women who had not received the vaccination or had one dose of vaccination before pregnancy, alive, known pregnant on the first day of follow-up, and aged 18 years or older. The exclusion criteria were women who were fully vaccinated (i.e. two vaccinations) before pregnancy, those for whom it was not possible to determine the start date of pregnancy due to unavailability of the gestational age and initial assessment dates in their records or those with miscarriage or stillbirth outcomes. Currently, within SAIL researchers are unable to account for terminations as these are classed as sensitive data and not currently accessible for research purposes.

### Calculating pregnancy start date

Pregnancy start dates were calculated from the following sources:

For pregnancies identified from the NCCH and MIDS data, the pregnancy start dates were calculated based on the gestational age and the week of birth data items available in these data sources. In cases where gestational age is missing, a value of 40 weeks was applied as the majority of those with missing data (92%) had birth weights suggestive of full-term infants. Thus, the pregnancy start date (last menstrual period) was simply calculated by subtracting the gestational age at birth (in weeks) from the week of birth. Pregnancies identified from both data sources were compared/matched and duplicate records were removed.

For pregnancies identified from the WLGP data, all pregnant women with a pregnancy code and event date that occurred during the study period were extracted (Supplementary Table 1). For those identified from the hospital admissions data (PEDW), all women with a pregnancy diagnosis code and an attendance date occurring during the study period were also extracted (Supplementary Table 2). Identified cases from both the WLGP and PEDW were separately matched to those identified from the NCCH and MIDS data to include only those who are still pregnant. Furthermore, the identified cases from both resources were further matched to remove duplicates and then linked to the initial assessment-related data items in the MIDS data. The gestational age in weeks and initial assessment data items are available in order to calculate the pregnancy start date. In cases where multiple records were found per pregnant woman, only the first occurring record between the study dates of interest was selected. The pregnancy start date for every successfully linked case was then calculated by subtracting the gestational age from the initial assessment date.

### Multimorbidity in pregnancy

Multimorbidity was defined by the presence of two or more long-term health conditions, which can include defined physical and mental health conditions [8]. Long-term health conditions are those that generally last a year or longer and have a significant impact on a person’s life [9]. Four long-term health conditions including depression, diabetes, asthma, and cardiovascular were selected on the following basis: (1) prevalence; (2) potential to impact vaccine uptake; and (3) recorded in the study datasets. These conditions were aggregated to generate a new multimorbidity variable with two distinct categories: Multimorbid and Non-multimorbid. The Multimorbid category comprises those with two or more health conditions, while the Non-multimorbid comprises healthy individuals together with those with only one health condition. Read codes for depression, diabetes, asthma, and cardiovascular can be found in Supplementary Tables 3 to 6 respectively. ICD-10 codes for the same conditions can be found in Supplementary Tables 7 to 10 respectively.

### Statistical analysis

Kaplan-Meier survival analysis was employed to examine time to vaccination by depression, diabetes, asthma, and cardiovascular diseases independently, by multimorbidity, as well as by smoking status censored at the birth, death, or moved out of Wales while pregnant. The Log Rank test was used to determine if there were differences in the survival distributions of vaccine uptake times within the diseases independently, multimorbidity, and smoking status. Differences were reported in median times (MD) with 95% confidence intervals and significance level accepted at p<0.05. Multivariate Cox regression hazard models were utilised to examine the impact of depression, diabetes, asthma, and cardiovascular diseases on vaccine acceptance independently with age group, ethnic group, area of deprivation, and smoking status incorporated into the model, as well as the impact of multimorbidity, age group, ethnic group, area of deprivation, and smoking status on vaccine acceptance, reporting hazard ratios (HR) with 95% confidence intervals and significance level accepted at p<0.05. Bootstrapping internal validation was conducted to assess the performance of the model, reporting bootstrapped Beta coefficients (B), standard error, 95% confidence intervals, and significance level accepted at p<0.05. The reference groups were those without multimorbidity, never smokers, aged 25-29, white ethnic group, and those living in the most affluent area. The data handling and preparation for the descriptive statistics, survival analysis, and Cox proportional hazard modelling were performed in an SQL IBM DB2 database within the SAIL Databank utilising Eclipse software. Final data preparation specific to these analyses, such as setting the reference groups was performed in IBM SPSS Statistics 28. Descriptive statistics, Survival, and Cox regression analyses were performed in SPSS.

## Results

A total of 28,343 pregnant women were identified from 13^th^ April 2021 through 31^st^ December 2021. After excluding women who were fully vaccinated before pregnancy (n=3,232), the cohort comprised 25,111 pregnant women. Those women were followed up, and their records were linked to the COVID-19 vaccination data up to and including 31^st^ December 2021. Over the study period, 8,203 (32.7%) of pregnant women received at least one dose of the COVID-19 vaccine during pregnancy, 8,572 (34.1%) were not vaccinated, and 8,336 (33.2%) received the vaccine after giving birth (Figure 1 describes the participants in the cohort). Most of the women were aged between 30-39, and between 25-29 years (48.4% and 29.7% respectively). The majority were White (77.8%). Nearly a quarter were living in the most deprived quintile (23.3%) and 14.4% were in the least deprived quintile. 29.5% of women were diagnosed with depression, 5.8% had diabetes, 23.9% had Asthma, 3.3% had cardiovascular, and 12.9% had two or more health conditions (Table 1).

**Table 1.** Descriptive summaries of the pregnant women eligible for vaccination

|  |  | N | % |
| --- | --- | --- | --- |
| Age group | 18-24 | 4,664 | 18.6 |
|  | 25-29 | 7,447 | 29.7 |
|  | 30-39 | 12,143 | 48.4 |
|  | 40-50 | 857 | 3.4 |
| Ethnic group | White<sup>1</sup> | 19,547 | 77.8 |
|  | Asian<sup>2</sup> | 902 | 3.6 |
|  | Mixed<sup>3</sup> | 316 | 1.3 |
|  | Black<sup>4</sup> | 440 | 1.8 |
|  | Other<sup>5</sup> | 571 | 2.3 |
|  | Unknown | 3,335 | 13.3 |
| WIMD Quintile 2019 | 1<sup>st</sup> (Most deprived) | 5,840 | 23.3 |
|  | 2<sup>nd</sup> | 4,795 | 19.1 |
|  | 3<sup>rd</sup> | 4,157 | 16.6 |
|  | 4<sup>th</sup> | 3,804 | 15.1 |
|  | 5<sup>th</sup> (Least deprived) | 3,626 | 14.4 |
|  | Unknown | 2,889 | 11.5 |
| **Smoking Status** | Current smoker | 4,441 | 17.1 |
|  | Former smoker | 3,150 | 12.5 |
|  | Never smoker | 12,418 | 49.5 |
|  | Unknown | 5,102 | 20.3 |
| **Depression** | Yes | 7,419 | 29.5 |
|  | No | 17,692 | 70.5 |
| **Diabetes** | Yes | 1,468 | 5.8 |
|  | No | 23,643 | 94.2 |
| **Asthma** | Yes | 5,999 | 23.9 |
|  | No | 19,112 | 76.1 |
| **Cardiovascular** | Yes | 826 | 3.3 |
|  | No | 24,285 | 96.7 |
| **Multimorbidity** | Yes | 3,247 | 12.9 |
|  | No | 21,864 | 87.1 |

<sup>1</sup> Comprises of Any White Background, Gypsy or Irish Traveller.
<sup>2</sup> Comprises of Bangladeshi, Pakistani, Indian, Any Other Asian Background.
<sup>3</sup> Comprises of Any Other Mixed Background, White and Asian, White and Black African, White and Black Caribbean, Any Other Mixed/Multiple Ethnic Background.
<sup>4</sup> Comprises of African, Any Other Black Background, Caribbean
<sup>5</sup> Comprises of Any Other Ethnic Group, Arab, Chinese.

**Fig. 1.**
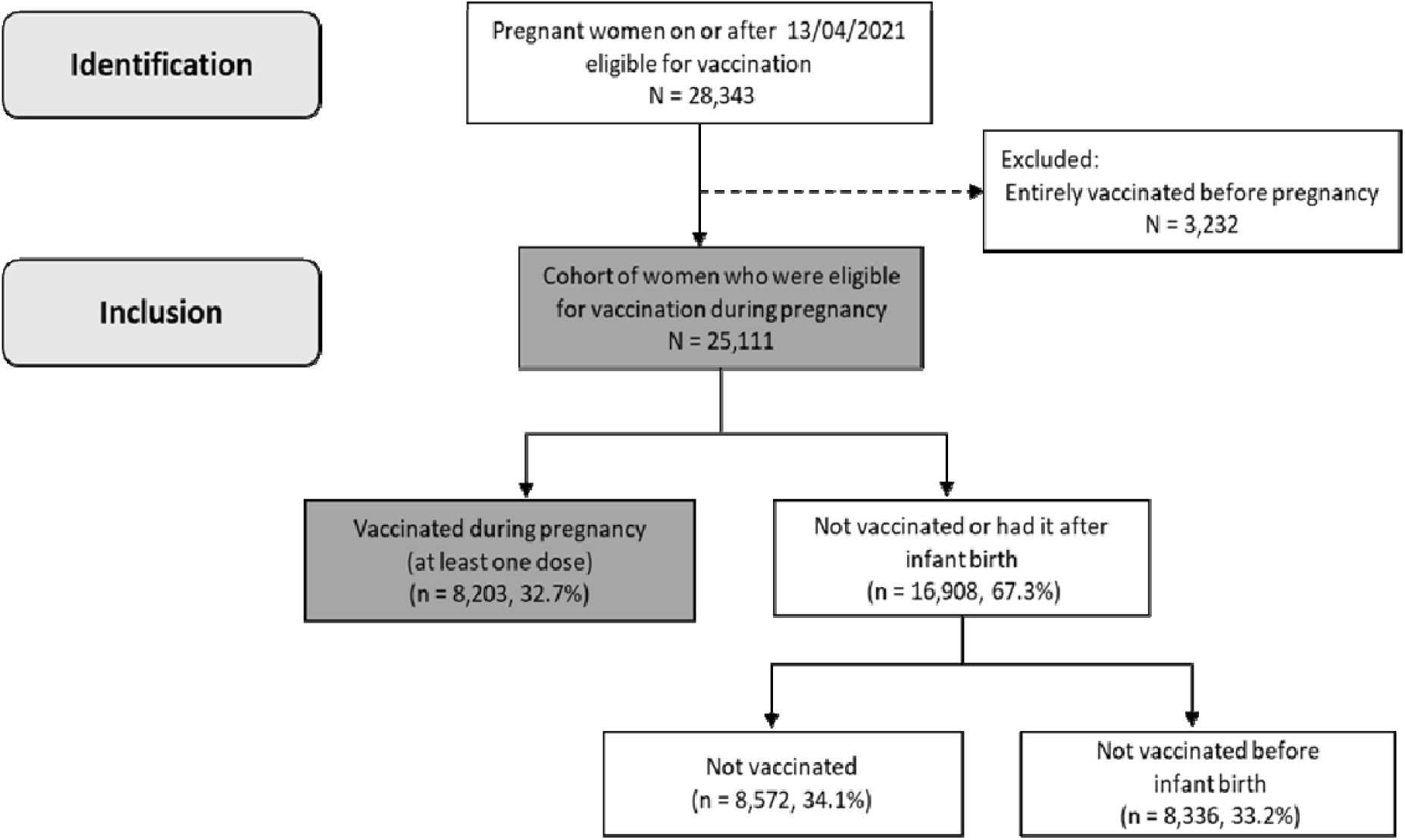
Flowchart of the cohort identification.

### Examining time to vaccination in pregnancy

Kaplan-Meier survival analysis indicates that pregnant women living with multimorbidity had a median time to vaccine uptake of 114 days (95% CI 106.6 to 121.4). This was lower than those with absence of multimorbidity, which had a median time to vaccine uptake of 126 days (95% CI 123.4 to 128.6). A Log Rank test was conducted to determine if there were significant differences in the survival distributions of vaccine uptake times for the different groups. The survival distributions were statistically significantly different, X^2^(1) = 5.4, p=0.02 (Figure 2a). However, other survival analyses were also conducted to estimate the time to vaccination for depression, diabetes, asthma, and cardiovascular diseases independently. The survival distributions were not significantly different with a significance level >0.05 between the different groups for all the diseases (Supplementary Table 11). Current Smoker women had a median time to vaccine uptake of 142 days (95% CI 132.6 to 151.4). This was longer than Never Smokers and Former Smokers, which had median times to vaccine uptake of 124 days (95% CI 120.9 to 127.1) and 129 days (95% CI 121.7 to 136.3) respectively. The survival distributions between smoking status groups were significantly different, X^2^(3) =40.23, p<.001. These differences were significant between the Current Smoker vs. the Never Smoker groups, X^2^(1) = 18, p<.001, and the Former Smoker vs. the Never Smoker group, X^2^(1)=4.74, p=0.03. However, the survival distribution for groups Current Smoker vs. Former Smoker was not significantly different, X^2^(1)=2.43, p=0.12 (Figure 2b).

**Fig 2a.**
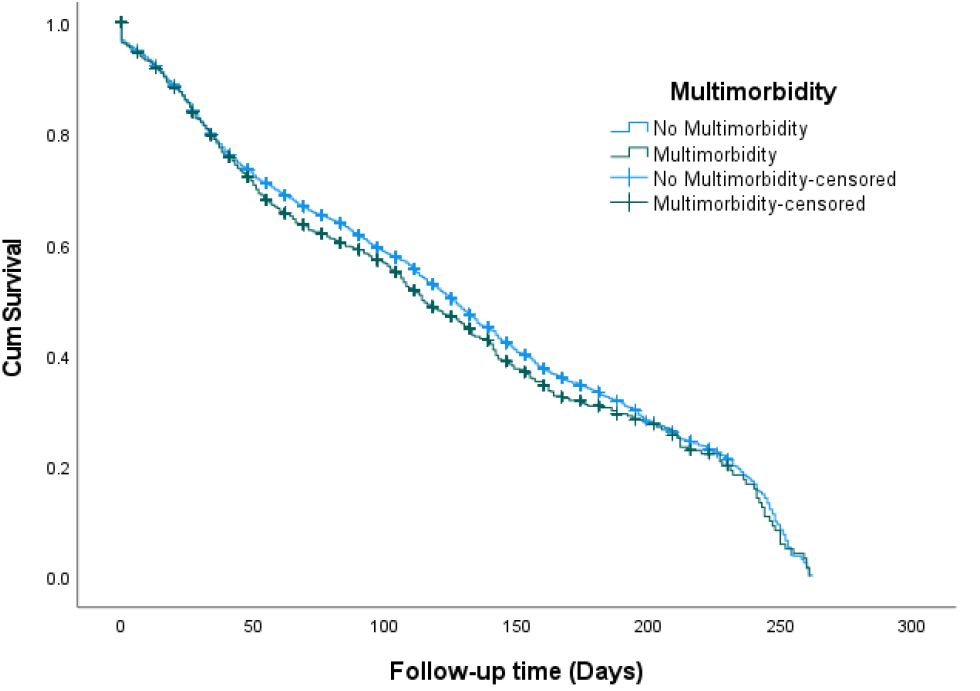
Time to vaccine uptake by Multimorbidity

**Fig 2b.**
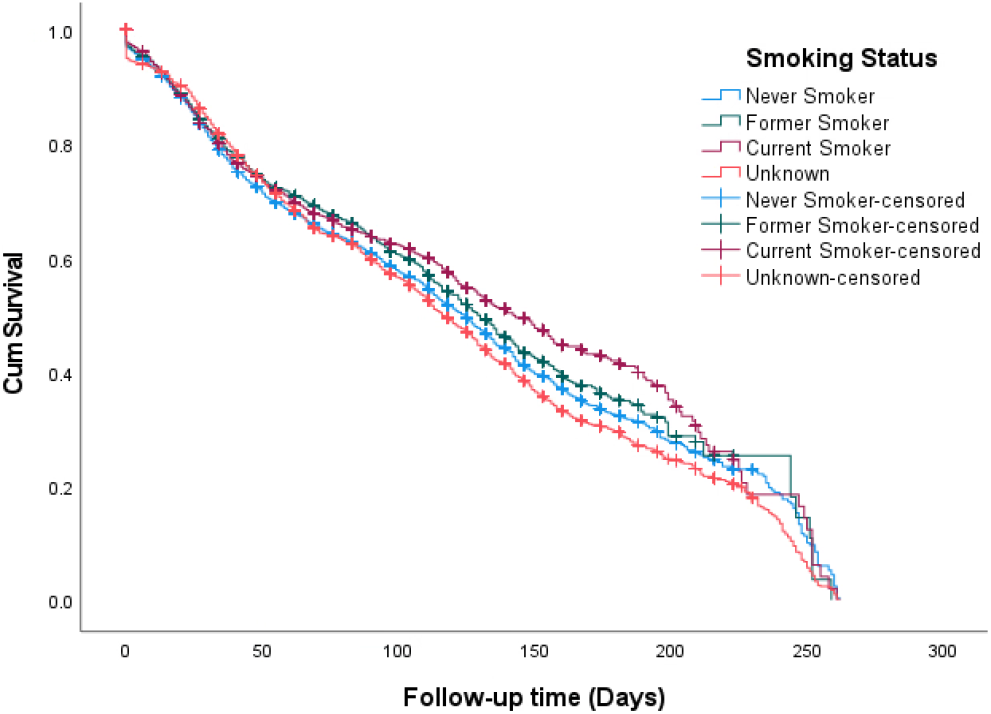
Time to vaccine uptake by Smoking status

### Examining the impact of multimorbidity, smoking status, and demographics on vaccine uptake

Women who had depression were slightly but significantly more likely to have the vaccine compared to those without depression (HR=1.08, 95% CI 1.03 to 1.14, p=.002) (supplementary table 12). Diabetes, asthma, and cardiovascular diseases were not associated with vaccine uptakes (p>0.05 for all) (Supplementary Tables 13-15). Results indicate that women living with multimorbidity were 1.12 times more likely to have the vaccine compared to those with no multimorbidity (HR=1.12, 95% CI 1.04 to 1.19, p=.001). Vaccine uptakes were significantly lower among both Current Smokers and Former Smokers compared to Never Smokers (HR=.87, 95% CI .81 to .94, p<.001) and (HR=.92, 95% CI .85 to .98, p=.015) respectively. Those aged 40-50 were 1.33 times more likely to have the vaccine compared to those aged 25-29 (HR=1.33, 95% CI 1.18 to 1.49, p<.001), also those aged 30-39 were 1.17 times more likely to have the vaccine compared to those aged 25-29 (HR=1.17, 95% CI 1.12 to 1.24, p<.001). It was also observed that the vaccine uptake was lower among those living in the most deprived areas compared to those living in the most affluent areas (HR=.89, 95% CI 0.83 to 0.96, p=.002). (Table 2).

**Table 2.** Cox Regression analysis of factors associated with vaccination uptake among pregnant women eligible for vaccination, adjusted analysis.

| Characteristic |  | HR<sup>1</sup> (95% CI<sup>2</sup>) | P value<sup>3</sup> |
| --- | --- | --- | --- |
| Age group | 25-29 | Reference |  |
|  | 18-24 | .99 (0.92 – 1.07) | .788 |
|  | 30-39 | 1.17 (1.12 – 1.24) | <.001 |
|  | 40-50 | 1.33 (1.18 – 1.49) | <.001 |
| Ethnic groups |  |  |  |
|  | White | Reference |  |
|  | Asian | 1.09 (.97 – 1.22) | .140 |
|  | Mixed | 1.02 (.81 – 1.28) | .899 |
|  | Black | 1.06 (.87 – 1.29) | .577 |
|  | Other | 1.16 (1.00 – 1.33) | .050 |
|  | Unknown | .94 (.87 – 1.01) | .067 |
| **WIMD quintile 2019** | 5<sup>th</sup> (Least deprived) | Reference |  |
|  | 4<sup>th</sup> | .90 (.84 – .97) | .005 |
|  | 3<sup>rd</sup> | .82 (.76 – .88) | <.001 |
|  | 2<sup>nd</sup> | .92 (0.85 – 0.99) | .018 |
|  | 1<sup>st</sup> (Most deprived) | .89 (0.83 – 0.96) | .002 |
| **Smoking status** | Never Smoker | Reference |  |
|  | Former Smoker | .92 (.85 - .98) | .015 |
|  | Current Smoker | .87 (.81 - .94) | <.001 |
|  | Unknown | 1.09 (1.03 - 1.15) | .003 |
| **Multimorbidity** | No Multimorbidity | Reference |  |
|  | Multimorbidity | 1.12 (1.04 - 1.19) | .001 |

<sup>1</sup>Hazard Ratio, <sup>2</sup>Confidence Interval (95%), <sup>3</sup>significance level accepted at <0.05

## Internal validation

The model was internally validated to estimate its performance more accurately. Bootstrap resampling started with fitting the regression model in a bootstrap sample of 1000 random samples, which were generated with replacement from the original dataset. Bootstrapping analyses were performed on each random sample, and beta coefficient, standard error, and 95% Bias corrected accelerated (BCa) confidence intervals for the primary findings were generated. Bootstrapping estimated the internal validity, providing stable estimates with low bootstrapped bias, low standard errors, and robust confidence intervals for both the multimorbidity and depression models (Table 3, supplementary table 16 respectively).

**Table 3.**
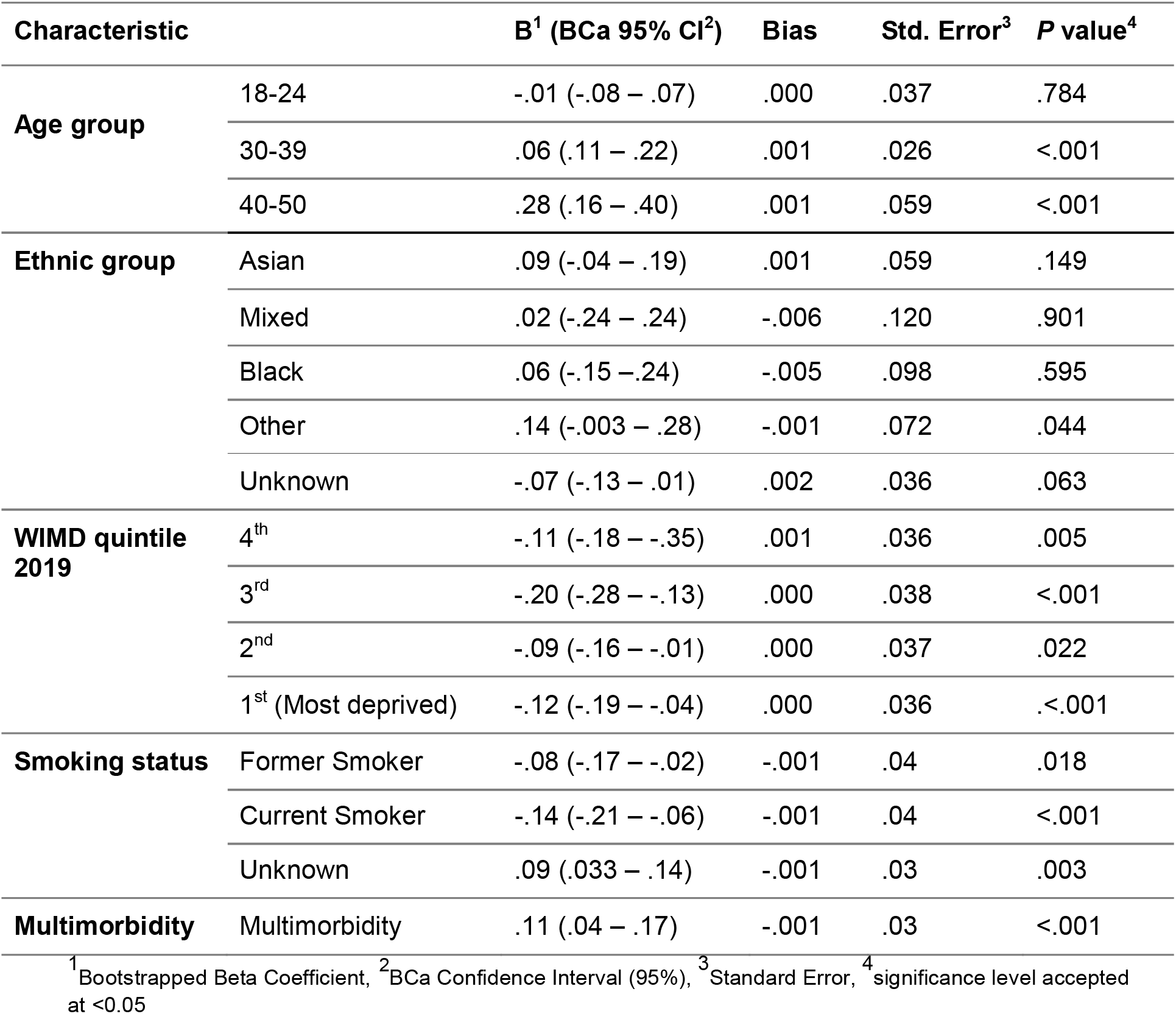
Bootstrapping internal validation of factors associated with vaccination uptake among pregnant women eligible for vaccination

## Discussion

This study investigated the impact of multimorbidity and smoking status on vaccine uptake as well as the impact of depression, diabetes, asthma, and cardiovascular diseases independently during pregnancy in Wales. Women with depression were slightly but significantly more likely to have the vaccine compared to those without depression. Those living with multimorbidity were also more likely to have the vaccine compared to those living with the absence of multimorbidity. Vaccine uptake was lower among those who currently smoke and those former smokers compared to those who have never smoked. Women aged 30 or older were more likely to have the vaccine compared to younger women. Uptake was lower among those living in the most deprived areas compared to those living in the most affluent areas. These findings may help to generate and tailor vaccine strategies to the populations who are more vaccine hesitant. The findings of the study complement previous studies of vaccine hesitancy. The presence of one or more chronic conditions has been found to be associated with the willingness to receive the COVID-19 vaccine [11]; this study supported that those with multimorbidity are more likely to accept the vaccine than non-multimorbid individuals. In previous studies, current smokers reported significantly greater mistrust of vaccines and any benefits and were more worried about future outcomes [13]. Current smokers were more likely to be unwilling to have the vaccine; 21.5% compared to 11.6% of never smokers, (OR=2.12, 95% CI 1.91-2.34, p<.001) and compared to 14.7% of former smokers (OR=1.53, 95% CI 1.37-1.71, p<.001) [13]. The findings of the study also indicated that current and former smokers were less likely to accept the COVID-19 vaccine. The ‘Understanding Society’ COVID-19 survey asked participants (n = 12,035) their likelihood of vaccine uptake and reason for hesitancy. Vaccine hesitancy was high in Black (71.8%) (OR= 13.42, 95% CI 6.86-26.24, p=<.05) and Pakistani/Bangladeshi (42.3%) OR=2.54, 95% CI 1.19-5.44, p=<.05 ethnic groups (compared to White British/Irish) [15]. The results indicate that those living in more deprived areas in Wales were less likely to accept the vaccine. Similar results were found with adults living in the most deprived areas of England (based on the Index of Multiple Deprivation) were more likely to report vaccine hesitancy (8%) than adults living in the least deprived areas (2%) [16].

## Strengths and limitations

The study has several strengths; it utilises primary and secondary health care data for pregnant women in Wales including the maternity and child health data, it gives a national perspective of COVID-19 vaccine hesitancy, making the findings generalizable due to its total population cohort. Bootstrapping internal validation was performed to estimate the performance of the models with increased accuracy. The study had some limitations, such as not indicating in which trimester pregnant women had the vaccine: it has been reported that pregnant women in the first trimester expressed higher acceptance of COVID-19 vaccination than those in the second and third trimesters [14]. The study excluded miscarriage and stillbirth outcomes as these are classed by SAIL as sensitive data and are not currently accessible for research purposes.

## Conclusion

In conclusion, it is critical to develop tailored strategies to increase the acceptance rates of the COVID-19 vaccine and decrease hesitancy. A more targeted approach to vaccinations may need to be addressed to reach certain groups such as younger people, smokers, healthy individuals, and those living in higher deprivation level areas. Encouraging vulnerable populations including pregnant women is a priority moving forward.

## Data Availability

The data that support the findings of this study are available from SAIL, but restrictions apply to the availability of these data, which were used under license for the current study, and so are not publicly available. Data are however available from the authors upon reasonable request and with permission of SAIL.

## List of abbreviations

EHR: Electronic Health Records
SAIL: Secure Anonymised Information Linkage
HR: Hazard Ratios
SAGE: The Scientific Advisory Group for Emergencies
JCVI: The UK’s Joint Committee on Vaccination and Immunisation
GP: General Practitioners
WLGP: Wales Longitudinal General Practice
PEDW: Patient Episode Database for Wales
OPDW: Outpatient Database for Wales
NCCH: National Community Child Health
MIDS: Maternal Indicators Dataset
CVVD: COVID-19 Vaccination Data
WDSD: Welsh Demographic Service Dataset
WIMD: Welsh Index for Multiple Deprivation
MD: Median Times
SQL: Structured Query Language
SPSS: Statistical Package for Social Sciences
WHO: The World Health Organisation
IGRP: Information Governance Review Panel
HDR: Health Data Research
BHF: British Heart Foundation
UK: United Kingdom

## Declarations

### Ethical approval and consent to participate

The data held in the SAIL (Secure Anonymised Information Linkage) Databank in Wales/UK are anonymised. All data contained in SAIL has the permission from the relevant Caldicott Guardian or Data Protection Officer and SAIL-related projects are required to obtain Information Governance Review Panel (IGRP) approval. The IGRP approval number for this study is 0911. The Research Ethics Service approval was also given by the North West Greater Manchester East Research Ethics Committee for the qualitative research. The Ethics Reference Number is RIO 030-20.

### Consent for publication

Not applicable

### Authors’ Contributions

SB, MM, HJ, and JK conceived the article. MM carried out the quantitative analysis. MM and HJ prepared the first draft of the manuscript. MS, MP, NK, AA, LZ, ALA, AD, KN contributed substantially to the manuscript, writing - reviewing & editing. All authors approved the final version submitted for publication

## Acknowledgments

This study is part of the National Centre for Population Health and Wellbeing, which is funded by Health Care Research Wales. This study makes use of anonymised data held in the Secure Anonymised Information Linkage (SAIL) Databank (17, 20, 21). We would like to acknowledge all the data providers who make anonymised data available for research.

This work was supported by Health Data Research UK, which receives its funding from HDR UK Ltd (HDR-9006) funded by the UK Medical Research Council, Engineering and Physical Sciences Research Council, Economic and Social Research Council, Department of Health and Social Care (England), Chief Scientist Office of the Scottish Government Health and Social Care Directorates, Health and Social Care Research and Development Division (Welsh Government), Public Health Agency (Northern Ireland), British Heart Foundation (BHF) and the Welcome Trust. This work was supported by the ADR Wales programme of work. The ADR Wales programme of work is aligned to the priority themes as identified in the Welsh Government’s national strategy: Prosperity for All. ADR Wales brings together data science experts at Swansea University Medical School, staff from the Wales Institute of Social and Economic Research, Data and Methods (WISERD) at Cardiff University and specialist teams within the Welsh Government to develop new evidence which supports Prosperity for All by using the SAIL Databank at Swansea University, to link and analyse anonymised data. ADR Wales is part of the Economic and Social Research Council (part of UK Research and Innovation) funded ADR UK (grant ES/S007393/1).

This work was additionally supported by funding from the Data and Connectivity National Core Study, led by Health Data Research UK in partnership with the Office for National Statistics and funded by UK Research and Innovation (grant ref MC_PC_20058), with additional support by The Alan Turing Institute via ‘Towards Turing 2.0’ EPSRC Grant Funding.

The responsibility for the interpretation of the information supplied is the authors’ alone.

## Additional files

Supplementary Tables - Impact of Multimorbidity on COVID-19 Vaccine Hesitancy

