## Supplemental File for "COVID-19 Vaccination in Pregnancy: The Impact of Multimorbidity and Smoking Status on Vaccine Hesitancy, a Cohort Study of 25,111 Women in Wales, UK"

Supplementary table 1: Read codes (v2) used to identify pregnancy from the primary care data (GP)

| **Read code** | **Description** |
| --- | --- |
| 13H7. | Unwanted pregnancy |
| 2711. | O/E - fundus 12-16 week size |
| 2712. | O/E - fundus 16-20 week size |
| 2713. | O/E - fundus 20-24 week size |
| 2714. | O/E - fundus 24-28 week size |
| 2715. | O/E - fundus 28-32 week size |
| 2717. | O/E - fundus 34-36 week size |
| 4453. | Serum pregnancy test positive |
| 4654. | Urine pregnancy test positive |
| 584.. | Ultrasound in obstetric diagnosis |
| 5841. | U-S obstetric scan requested |
| 5842. | U-S obstetric scan normal |
| 584B. | Viability US scan |
| 584C. | Antenatal ultrasound result received |
| 584D. | Antenatal ultrasound confirms intra-uterine pregnancy |
| 584Z. | U-S obstetric diagnosis scan NOS |
| 62... | Patient pregnant |
| 621.. | Patient currently pregnant |
| 6212. | Pregnant - blood test confirms |
| 6214. | Pregnant - on history |
| 6216. | Pregnant - planned |
| 6217. | Pregnant - unplanned - wanted |
| 621C. | Unplanned pregnancy |
| 621Z. | Patient pregnant NOS |
| 622.. | Antenatal care: gravida No. |
| 623.. | A/N care: obstetric risk |
| 625Z. | A/N care: social risk NOS |
| 628Z. | A/N risk NOS |
| 62A.. | A/N care provider |
| 62A3. | A/N - shared care |
| 62B.. | Delivery booking place |
| 62B3. | G.P. unit delivery booking |
| 62B4. | Consultant unit booking |
| 62B8. | Midwife unit delivery booking |
| 62C.. | Delivery booking - length of stay |
| 62F.. | Antenatal amniocentesis |
| 62G.. | Antenatal ultrasound scan |
| 62GB. | Antenatal ultrasounds scan at 4-8 weeks |
| 62GZ. | Antenatal ultrasound scan NOS |
| 62L.. | Antenatal blood group screen |
| 62N.. | Antenatal examinations |
| 62N1. | A/N booking examination |
| 62N3. | A/N 16-week examination |
| 62O1. | Fetal movements felt |
| 62X.. | Length of gestation |
| 62Y.. | Routine antenatal care |
| 62a.. | Pregnancy review |
| 62b.. | Antenatal HIV screening |
| 62c.. | Antenatal screening |
| 6776. | Pregnancy termination counselling |
| 679E. | Antenatal education |
| 7F051 | Diagnostic amniocentesis |
| 7F060 | Cerclage of cervix of gravid uterus |
| 7F25. | Obstetric monitoring |
| 7F2B1 | Ultrasound monitoring of early pregnancy |
| 8B75. | Vitamin supplement - pregnancy |
| 8H7W. | Refer to TOP counselling |
| 8HHV. | Referral for termination of pregnancy |
| 8HHf. | Refer to early pregnancy unit |
| 8HT9. | Referral to antenatal clinic |
| 8M6.. | Requests pregnancy termination |
| 95... | Maternity services admin. |
| 9N1N. | Seen in antenatal clinic |
| L1... | Pregnancy complications |
| L10.. | Haemorrhage in early pregnancy |
| L100. | Threatened abortion |
| L10y. | Other haemorrhage in early pregnancy |
| L11.. | Antepartum haemorrhage, abruptio placentae, placenta praevia |
| L1246 | Pre-eclampsia, unspecified |
| L13.. | Excessive pregnancy vomiting |
| L130. | Mild hyperemesis gravidarum |
| L1300 | Mild hyperemesis unspecified |
| L13y. | Other pregnancy vomiting |
| L13z. | Unspecified pregnancy vomiting |
| L1668 | Urinary tract infection complicating pregnancy |
| L16y5 | Abdominal pain in pregnancy |
| L1808 | Diabetes mellitus arising in pregnancy |
| L1809 | Gestational diabetes mellitus |
| L1825 | Iron deficiency anaemia of pregnancy |
| L18A0 | Cholestasis of pregnancy |
| L210. | Twin pregnancy |
| L25.. | Known or suspected fetal abnormality |
| L264. | Intrauterine death |
| L280. | Oligohydramnios |
| L33z. | Umbilical cord complications NOS |
| L413. | Antenatal deep vein thrombosis |
| L510. | Maternal care for hydrops fetalis |
| Lyu21 | [X]Other vomiting complicating pregnancy |
| Lyu25 | [X]Other specified pregnancy-related conditions |
| Z212. | Antenatal care |
| Z22.. | Pregnancy observations |
| Z225. | Normal pregnancy |
| Z226. | Pregnancy problem |
| Z227. | Confirmation of pregnancy |
| Z2291 | Intrauterine pregnancy |
| Z22A. | Observation of pattern of pregnancy |
| Z22A1 | Low risk pregnancy |
| Z22A4 | Early stage of pregnancy |
| Z22AA | Wanted pregnancy |
| Z22AB | Unplanned pregnancy |
| Z22B1 | Single pregnancy |
| Z22C1 | Estimated date of delivery from last period |
| Z22C3 | Length of gestation |
| Z22D1 | Viable pregnancy |
| Z22D2 | Non-viable pregnancy |
| Z22D3 | Uncertain viability of pregnancy |
| ZV222 | [V]Pregnancy confirmed |
| ZV223 | [V]Pregnant state, incidental |
| ZV231 | [V]Pregnancy with history of trophoblastic disease |
| ZV28. | [V]Antenatal screening |
| 1531. | Last menstural period-1st day |
| 271B. | O/E - fundal size = dates |
| 2726. | O/E - fetal presentation unsure |
| 2766. | O/E - fetal heart 120-160 |
| 5391. | Obstetric X-ray - fetus |
| 584A. | Dating/Booking US scan |
| 67A.. | Pregnancy advice |
| 67A2. | Diet in pregnancy advice |
| 67A3. | Pregnancy smoking advice |
| 67AE. | Folic acid advice in first trimester of pregnancy |
| 67AF. | Pregnancy advice for patients with epilepsy |
| 7F261 | Viability scan |
| 7F2B. | Obstetric ultrasound monitoring |
| 9511. | FP24 signed by patient |
| 957.. | FW 8-applic for presc exempt |
| 9kv.. | Pertussis vaccination programme pregnant women enhance service admin |
| 9mK.. | Pertussis vaccination in pregnancy invitation |
| 9Nk3. | Seen in fetal medicine clinic |
| 9NkN. | Seen in early pregnancy unit |
| 9NV1. | Antenatal clinic |
| ZV286 | [V] Antenatal screening for chromosomal anomalies |
| 6556. | Pretussis vaccination in pregnancy |

Supplementary table 2: ICD-10 version:2019 codes used to identify pregnancy from the hospital admissions data (PEDW)

| **ICD-10 code** | **Definition** |
| --- | --- |
| O11X | Pre-eclampsia superimposed on chronic hypertension |
| O120 | Gestational oedema |
| O121 | Gestational proteinuria |
| O122 | Gestational oedema with proteinuria |
| O13X | Gestational [pregnancy-induced] hypertension |
| O140 | Mild to moderate pre-eclampsia |
| O141 | Severe pre-eclampsia |
| O142 | HELLP syndrome |
| O149 | Pre-eclampsia, unspecified |
| O150 | Eclampsia in pregnancy |
| O116X | Unspecified maternal hypertension |
| O200 | Threatened abortion |
| O208 | Other haemorrhage in early pregnancy |
| O209 | Haemorrhage in early pregnancy, unspecified |
| O20 | Haemorrhage in early pregnancy |
| O210 | Mild hyperemesis gravidarum |
| O211 | Hyperemesis gravidarum with metabolic disturbance |
| O212 | Late vomiting of pregnancy |
| O219 | Vomiting of pregnancy, unspecified |
| O220 | Varicose veins of lower extremity in pregnancy |
| O223 | Deep phlebothrombosis in pregnancy |
| O224 | Haemorrhoids in pregnancy |
| O228 | Other venous complications in pregnancy |
| O229 | Venous complication in pregnancy, unspecified |
| O230 | Infections of kidney in pregnancy |
| O234 | Unspecified infection of urinary tract in pregnancy |
| O235 | Infections of the genital tract in pregnancy |
| O239 | Other and unspecified genitourinary tract infection in pregnancy |
| O244 | Diabetes mellitus arising in pregnancy |
| O249 | Diabetes mellitus in pregnancy, unspecified |
| O260 | Excessive weight gain in pregnancy |
| O261 | Low weight gain in pregnancy |
| O262 | Pregnancy care of habitual aborter |
| O265 | Maternal hypotension syndrome |
| O268 | Other specified pregnancy-related conditions |
| O269 | Pregnancy-related condition, unspecified |
| O280 | Abnormal haematological finding on antenatal screening of mother |
| O281 | Abnormal biochemical finding on antenatal screening of mother |
| O283 | Abnormal ultrasonic finding on antenatal screening of mother |
| O289 | Abnormal finding on antenatal screening of mother, unspecified |
| O300 | Twin pregnancy |
| O301 | Triplet pregnancy |
| O320 | Maternal care for unstable lie |
| O321 | Maternal care for breech presentation |
| O322 | Maternal care for transverse and oblique lie |
| O324 | Maternal care for high head at term |
| O326 | Maternal care for compound presentation |
| O328 | Maternal care for other malpresentation of fetus |
| O329 | Maternal care for malpresentation of fetus, unspecified |
| O340 | Maternal care for congenital malformation of uterus |
| O341 | Maternal care for tumour of corpus uteri |
| O342 | Maternal care due to uterine scar from previous surgery |
| O343 | Maternal care for cervical incompetence |
| O344 | Maternal care for other abnormalities of cervix |
| O346 | Maternal care for abnormality of vagina |
| O347 | Maternal care for abnormality of vulva and perineum |
| O348 | Maternal care for other abnormalities of pelvic organs |
| O350 | Maternal care for (suspected) central nervous system malformation in fetus |
| O351 | Maternal care for (suspected) chromosomal abnormality in fetus |
| O352 | Maternal care for (suspected) hereditary disease in fetus |
| O358 | Maternal care for other (suspected) fetal abnormality and damage |
| O359 | Maternal care for (suspected) fetal abnormality and damage, unspecified |
| O35 | Maternal care for known or suspected fetal abnormality and damage |
| O360 | Maternal care for rhesus isoimmunization |
| O361 | Maternal care for other isoimmunization |
| O363 | Maternal care for signs of fetal hypoxia |
| O364 | Maternal care for intrauterine death |
| O365 | Maternal care for poor fetal growth |
| O366 | Maternal care for excessive fetal growth |
| O368 | Maternal care for other specified fetal problems |
| O369 | Maternal care for fetal problem, unspecified |
| O40X | Polyhydramnios |
| O410 | Oligohydramnios |
| O418 | Other specified disorders of amniotic fluid and membranes |
| O429 | Premature rupture of membranes, unspecified |
| O438 | Other placental disorders |
| O440 | Placenta praevia specified as without haemorrhage |
| O441 | Placenta praevia with haemorrhage |
| O459 | Premature separation of placenta, unspecified |
| O468 | Other antepartum haemorrhage |
| O469 | Antepartum haemorrhage, unspecified |
| O470 | False labour before 37 completed weeks of gestation |
| O471 | False labour at or after 37 completed weeks of gestation |
| O479 | False labour, unspecified |
| O48X | Prolonged pregnancy |
| O718 | Other specified obstetric trauma |
| O882 | Obstetric blood-clot embolism |
| Z321 | Pregnancy confirmed |
| Z33X | Pregnant state, incidental |
| Z340 | Supervision of normal first pregnancy |
| Z348 | Supervision of other normal pregnancy |
| Z349 | Supervision of normal pregnancy, unspecified |
| Z352 | Supervision of pregnancy with other poor reproductive or obstetric history |
| Z353 | Supervision of pregnancy with history of insufficient antenatal care |
| Z357 | Supervision of high-risk pregnancy due to social problems |
| Z358 | Supervision of other high-risk pregnancies |
| Z368 | Other antenatal screening |
| Z369 | Antenatal screening, unspecified |

Supplementary table 3: Read codes (v2) used to identify depression from the primary care data (GP)

| Read code | Description |
| --- | --- |
| 1465 | H/O: depression |
| 212S. | Depression resolved |
| 8BK0. | Depression management programme |
| 8CAa. | Patient given advice about management of depression |
| 8HHq. | Referral for guided self-help for depression |
| 9H90. | Depression annual review |
| 9H91. | Depression medication review |
| 9H92. | Depression interim review |
| 9HA0. | On depression register |
| 9k40. | Depression - enhanced service completed |
| 9k4.. | Depression - enhanced services administration |
| 9kQ.. | On full dose long term treatment depression - enh serv admin |
| 9Ov0. | Depression monitoring first letter |
| 9Ov1. | Depression monitoring second letter |
| 9Ov2. | Depression monitoring third letter |
| 9Ov3. | Depression monitoring verbal invite |
| 9Ov4. | Depression monitoring telephone invite |
| 9Ov.. | Depression monitoring administration |
| E0013 | Presenile dementia with depression |
| E0021 | Senile dementia with depression |
| E0043 | Arteriosclerotic dementia with depression |
| E1120 | Single major depressive episode, unspecified |
| E1121 | Single major depressive episode, mild |
| E1122 | Single major depressive episode, moderate |
| E1123 | Single major depressive episode, severe, without psychosis |
| E1124 | Single major depressive episode, severe, with psychosis |
| E1125 | Single major depressive episode, partial or unspec remission |
| E1126 | Single major depressive episode, in full remission |
| E112. | Single major depressive episode |
| E112z | Single major depressive episode NOS |
| E1130 | Recurrent major depressive episodes, unspecified |
| E1131 | Recurrent major depressive episodes, mild |
| E1132 | Recurrent major depressive episodes, moderate |
| E1133 | Recurrent major depressive episodes, severe, no psychosis |
| E1134 | Recurrent major depressive episodes, severe, with psychosis |
| E1135 | Recurrent major depressive episodes,partial/unspec remission |
| E1136 | Recurrent major depressive episodes, in full remission |
| E1137 | Recurrent depression |
| E113. | Recurrent major depressive episode |
| E113z | Recurrent major depressive episode NOS |
| E118. | Seasonal affective disorder |
| E11.. | Depressive psychoses |
| E11y2 | Atypical depressive disorder |
| E11z2 | Masked depression |
| E130. | Reactive depressive psychosis |
| E135. | Agitated depression |
| E2003 | Anxiety with depression |
| E291. | Prolonged depressive reaction |
| E2B1. | Chronic depression |
| E2B.. | Depressive disorder NEC |
| Eu204 | [X]Post-schizophrenic depression |
| Eu251 | [X]Schizoaffective disorder, depressive type |
| Eu320 | [X]Mild depressive episode |
| Eu321 | [X]Moderate depressive episode |
| Eu322 | [X]Severe depressive episode without psychotic symptoms |
| Eu323 | [X]Severe depressive episode with psychotic symptoms |
| Eu324 | [X]Mild depression |
| Eu325 | [X]Major depression, mild |
| Eu326 | [X]Major depression, moderately severe |
| Eu327 | [X]Major depression, severe without psychotic symptoms |
| Eu328 | [X]Major depression, severe with psychotic symptoms |
| Eu329 | [X]Single major depr ep, severe with psych, psych in remiss |
| Eu32A | [X]Recurr major depr ep, severe with psych, psych in remiss |
| Eu32. | [X]Depressive episode |
| Eu32y | [X]Other depressive episodes |
| Eu32z | [X]Depressive episode, unspecified |
| Eu330 | [X]Recurrent depressive disorder, current episode mild |
| Eu331 | [X]Recurrent depressive disorder, current episode moderate |
| Eu332 | [X]Recurr depress disorder cur epi severe without psyc sympt |
| Eu333 | [X]Recurrent depress disorder cur epi severe with psyc symp |
| Eu334 | [X]Recurrent depressive disorder, currently in remission |
| Eu33. | [X]Recurrent depressive disorder |
| Eu33y | [X]Other recurrent depressive disorders |
| Eu33z | [X]Recurrent depressive disorder, unspecified |
| Eu341 | [X]Dysthymia |
| Eu412 | [X]Mixed anxiety and depressive disorder |

Supplementary table 4: Read codes (v2) used to identify diabetes from the primary care data (GP)

| **Read code** | **Description** |
| --- | --- |
| C1000 | Diabetes mellitus, juvenile type, no mention of complication |
| C1010 | Diabetes mellitus, juvenile type, with ketoacidosis |
| C1020 | Diabetes mellitus, juvenile type, with hyperosmolar coma |
| C1030 | Diabetes mellitus, juvenile type, with ketoacidotic coma |
| C1040 | Diabetes mellitus, juvenile type, with renal manifestation |
| C1050 | Diabetes mellitus, juvenile type, + ophthalmic manifestation |
| C1060 | Diabetes mellitus, juvenile, + neurological manifestation |
| C1070 | Diabetes mellitus, juvenile +peripheral circulatory disorder |
| C1073 | IDDM with peripheral circulatory disorder |
| C108. | Insulin dependent diabetes mellitus |
| C1080 | Insulin-dependent diabetes mellitus with renal complications |
| C1081 | Insulin-dependent diabetes mellitus with ophthalmic comps |
| C1082 | Insulin-dependent diabetes mellitus with neurological comps |
| C1083 | Insulin dependent diabetes mellitus with multiple complicatn |
| C1084 | Unstable insulin dependent diabetes mellitus |
| C1085 | Insulin dependent diabetes mellitus with ulcer |
| C1086 | Insulin dependent diabetes mellitus with gangrene |
| C1087 | Insulin dependent diabetes mellitus with retinopathy |
| C1088 | Insulin dependent diabetes mellitus - poor control |
| C1089 | Insulin dependent diabetes maturity onset |
| C108A | Insulin-dependent diabetes without complication |
| C108B | Insulin dependent diabetes mellitus with mononeuropathy |
| C108C | Insulin dependent diabetes mellitus with polyneuropathy |
| C108D | Insulin dependent diabetes mellitus with nephropathy |
| C108E | Insulin dependent diabetes mellitus with hypoglycaemic coma |
| C108F | Insulin dependent diabetes mellitus with diabetic cataract |
| C108G | Insulin dependent diab mell with peripheral angiopathy |
| C108H | Insulin dependent diabetes mellitus with arthropathy |
| C108J | Insulin dependent diab mell with neuropathic arthropathy |
| C10E. | Type 1 diabetes mellitus |
| C10E0 | Type 1 diabetes mellitus with renal complications |
| C10E1 | Type 1 diabetes mellitus with ophthalmic complications |
| C10E2 | Type 1 diabetes mellitus with neurological complications |
| C10E3 | Type 1 diabetes mellitus with multiple complications |
| C10E4 | Unstable type 1 diabetes mellitus |
| C10E5 | Type 1 diabetes mellitus with ulcer |
| C10E6 | Type 1 diabetes mellitus with gangrene |
| C10E7 | Type 1 diabetes mellitus with retinopathy |
| C10E8 | Type 1 diabetes mellitus - poor control |
| C10E9 | Type 1 diabetes mellitus maturity onset |
| C10EA | Type 1 diabetes mellitus without complication |
| C10EB | Type 1 diabetes mellitus with mononeuropathy |
| C10EC | Type 1 diabetes mellitus with polyneuropathy |
| C10ED | Type 1 diabetes mellitus with nephropathy |
| C10EE | Type 1 diabetes mellitus with hypoglycaemic coma |
| C10EF | Type 1 diabetes mellitus with diabetic cataract |
| C10EG | Type 1 diabetes mellitus with peripheral angiopathy |
| C10EH | Type 1 diabetes mellitus with arthropathy |
| C10EJ | Type 1 diabetes mellitus with neuropathic arthropathy |
| C10EK | Type 1 diabetes mellitus with persistent proteinuria |
| C10EL | Type 1 diabetes mellitus with persistent microalbuminuria |
| C10EM | Type 1 diabetes mellitus with ketoacidosis |
| C10EN | Type 1 diabetes mellitus with ketoacidotic coma |
| C10EP | Type 1 diabetes mellitus with exudative maculopathy |
| C10EQ | Type 1 diabetes mellitus with gastroparesis |
| C10ER | Latent autoimmune diabetes mellitus in adult |
| C10y0 | Diabetes mellitus, juvenile, + other speciﬁed manifestation |
| C10z0 | Diabetes mellitus, juvenile type, + unspeciﬁed complication |
| L1805 | Pre-existing diabetes mellitus, insulin-dependent |
| C1001 | Diabetes mellitus, adult onset, no mention of complication |
| C1011 | Diabetes mellitus, adult onset, with ketoacidosis |
| C1021 | Diabetes mellitus, adult onset, with hyperosmolar coma |
| C1031 | Diabetes mellitus, adult onset, with ketoacidotic coma |
| C1041 | Diabetes mellitus, adult onset, with renal manifestation |
| C1051 | Diabetes mellitus, adult onset, + ophthalmic manifestation |
| C1061 | Diabetes mellitus, adult onset, + neurological manifestation |
| C1071 | Diabetes mellitus, adult, + peripheral circulatory disorder |
| C1072 | Diabetes mellitus, adult with gangrene |
| C1074 | NIDDM with peripheral circulatory disorder |
| C109. | Non-insulin dependent diabetes mellitus |
| C1090 | Non-insulin-dependent diabetes mellitus with renal comps |
| C1091 | Non-insulin-dependent diabetes mellitus with ophthalm comps |
| C1092 | Non-insulin-dependent diabetes mellitus with neuro comps |
| C1093 | Non-insulin-dependent diabetes mellitus with multiple comps |
| C1094 | Non-insulin dependent diabetes mellitus with ulcer |
| C1095 | Non-insulin dependent diabetes mellitus with gangrene |
| C1096 | Non-insulin-dependent diabetes mellitus with retinopathy |
| C1097 | Non-insulin dependent diabetes mellitus - poor control |
| C1099 | Non-insulin-dependent diabetes mellitus without complication |
| C109A | Non-insulin dependent diabetes mellitus with mononeuropathy |
| C109B | Non-insulin dependent diabetes mellitus with polyneuropathy |
| C109C | Non-insulin dependent diabetes mellitus with nephropathy |
| C109D | Non-insulin dependent diabetes mellitus with hypoglyca coma |
| C109E | Non-insulin depend diabetes mellitus with diabetic cataract |
| C109F | Non-insulin-dependent d m with peripheral angiopath |
| C109G | Non-insulin dependent diabetes mellitus with arthropathy |
| C109H | Non-insulin dependent d m with neuropathic arthropathy |
| C109J | Insulin treated Type 2 diabetes mellitus |
| C109K | Hyperosmolar non-ketotic state in type 2 diabetes mellitus |
| C10F. | Type 2 diabetes mellitus |
| C10F0 | Type 2 diabetes mellitus with renal complications |
| C10F1 | Type 2 diabetes mellitus with ophthalmic complications |
| C10F2 | Type 2 diabetes mellitus with neurological complications |
| C10F3 | Type 2 diabetes mellitus with multiple complications |
| C10F4 | Type 2 diabetes mellitus with ulcer |
| C10F5 | Type 2 diabetes mellitus with gangrene |
| C10F6 | Type 2 diabetes mellitus with retinopathy |
| C10F7 | Type 2 diabetes mellitus - poor control |
| C10F9 | Type 2 diabetes mellitus without complication |
| C10FA | Type 2 diabetes mellitus with mononeuropathy |
| C10FB | Type 2 diabetes mellitus with polyneuropathy |
| C10FC | Type 2 diabetes mellitus with nephropathy |
| C10FD | Type 2 diabetes mellitus with hypoglycaemic coma |
| C10FE | Type 2 diabetes mellitus with diabetic cataract |
| C10FF | Type 2 diabetes mellitus with peripheral angiopathy |
| C10FG | Type 2 diabetes mellitus with arthropathy |
| C10FH | Type 2 diabetes mellitus with neuropathic arthropathy |
| C10FJ | Insulin treated Type 2 diabetes mellitus |
| C10FK | Hyperosmolar non-ketotic state in type 2 diabetes mellitus |
| C10FL | Type 2 diabetes mellitus with persistent proteinuria |
| C10FM | Type 2 diabetes mellitus with persistent microalbuminuria |
| C10FN | Type 2 diabetes mellitus with ketoacidosis |
| C10FQ | Type 2 diabetes mellitus with exudative maculopathy |
| C10FR | Type 2 diabetes mellitus with gastroparesis |
| C10y1 | Diabetes mellitus, adult, + other speciﬁed manifestation |
| C10z1 | Diabetes mellitus, adult onset, + unspeciﬁed complication |
| L1806 | Pre-existing diabetes mellitus, non-insulin-dependent |
| C10.. | Diabetes mellitus |
| C100. | Diabetes mellitus with no mention of complication |
| C100z | Diabetes mellitus NOS with no mention of complication |
| C101. | Diabetes mellitus with ketoacidosis |
| C101y | Other speciﬁed diabetes mellitus with ketoacidosis |
| C101z | Diabetes mellitus NOS with ketoacidosis |
| C102. | Diabetes mellitus with hyperosmolar coma |
| C102z | Diabetes mellitus NOS with hyperosmolar coma |
| C103. | Diabetes mellitus with ketoacidotic coma |
| C103y | Other speciﬁed diabetes mellitus with coma |
| C103z | Diabetes mellitus NOS with ketoacidotic coma |
| C104. | Diabetes mellitus with renal manifestation |
| C104y | Other speciﬁed diabetes mellitus with renal complications |
| C104z | Diabetes mellitus with nephropathy NOS |
| C105. | Diabetes mellitus with ophthalmic manifestation |
| C105y | Other speciﬁed diabetes mellitus with ophthalmic complicatn |
| C105z | Diabetes mellitus NOS with ophthalmic manifestation |
| C107. | Diabetes mellitus with peripheral circulatory disorder |
| C107y | Other speciﬁed diabetes mellitus with periph circ comps |
| C107z | Diabetes mellitus NOS with peripheral circulatory disorder |
| C108y | Other speciﬁed diabetes mellitus with multiple comps |
| C108z | Unspeciﬁed diabetes mellitus with multiple complications |
| C10y. | Diabetes mellitus with other speciﬁed manifestation |
| C10yy | Other speciﬁed diabetes mellitus with other spec comps |
| C10yz | Diabetes mellitus NOS with other speciﬁed manifestation |
| C10z. | Diabetes mellitus with unspeciﬁed complication |
| C10zy | Other speciﬁed diabetes mellitus with unspeciﬁed comps |
| C10zz | Diabetes mellitus NOS with unspeciﬁed complication |
| L180X | Pre-existing diabetes mellitus, unspeciﬁed |
| L1808 | Diabetes mellitus arising in pregnancy |
| L1809 | Gestational diabetes mellitus |
| L180. | Diabetes mellitus during pregnancy/childbirth/puerperium |
| L1800 | Diabetes mellitus - unspec whether in pregnancy/puerperium |
| L1801 | Diabetes mellitus during pregnancy - baby delivered |
| L1802 | Diabetes mellitus in puerperium - baby delivered |
| L1803 | Diabetes mellitus during pregnancy - baby not yet delivered |
| L1804 | Diabetes mellitus in pueperium - baby previously delivered |
| L180z | Diabetes mellitus in pregnancy/childbirth/puerperium NOS |

Supplementary table 5: Read codes (v2) used to identify asthma from the primary care data (GP)

| **Read code** | **Description** |
| --- | --- |
| 8795 | Asthma control step 2 |
| H3300 | Extrinsic asthma without status asthmaticus |
| H331. | Late onset asthma |
| 679J. | Health education - asthma |
| H334. | Brittle asthma |
| 663d. | Emergency asthma admission since last appointment |
| 9OJ5. | Asthma monitor 2nd letter |
| 8B3j. | Asthma medication review |
| H33z1 | Asthma attack |
| 1782 | Asthma trigger - tobacco smoke |
| 663t. | Asthma causes daytime symptoms 1 to 2 times per month |
| 66YQ. | Asthma monitoring by nurse |
| 388t. | Royal College of Physicians asthma assessment |
| 663Q. | Asthma not limiting activities |
| 66YC. | Absent from work or school due to asthma |
| 663V2 | Moderate asthma |
| 66YK. | Asthma follow-up |
| H33.. | Bronchial asthma |
| H3301 | Extrinsic asthma with asthma attack |
| 663V. | Asthma severity |
| H33zz | Asthma NOS |
| 663q. | Asthma daytime symptoms |
| H331. | Intrinsic asthma |
| 9OJ2. | Refuses asthma monitoring |
| 663e0 | Asthma sometimes restricts exercise |
| H33z1 | Asthma attack NOS |
| 1785 | Asthma trigger - damp |
| 663y. | Number of asthma exacerbations in past year |
| 663N. | Asthma disturbing sleep |
| 663P1 | Asthma limits activities 1 to 2 times per week |
| 663N2 | Asthma disturbs sleep frequently |
| 663j. | Asthma - currently active |
| 9OJB. | Asthma monitorng invit SMS (short message servce) txt messge |
| 8797 | Asthma control step 4 |
| H3311 | Intrinsic asthma with asthma attack |
| 1780 | Aspirin induced asthma |
| 663W. | Asthma prophylactic medication used |
| 663.. | Asthma monitoring |
| H332. | Mixed asthma |
| 679J1 | Health education - structured asthma discussion |
| H33z. | Asthma unspecified |
| 1788 | Asthma trigger - cold air |
| 9OJ3. | Asthma monitor offer default |
| 663r. | Asthma causes night symptoms 1 to 2 times per month |
| 661N1 | Asthma self-management plan review |
| 66Y9. | Step up change in asthma management plan |
| 663O. | Asthma not disturbing sleep |
| 66Yz0 | Asthma management plan declined |
| 66YA. | Step down change in asthma management plan |
| 9NNX. | Under care of asthma specialist nurse |
| 663V1 | Mild asthma |
| H3300 | Hay fever with asthma |
| H3311 | Intrinsic asthma with status asthmaticus |
| 66Yq. | Asthma causes night time symptoms 1 to 2 times per week |
| 8794 | Asthma control step 1 |
| H33z2 | Late-onset asthma |
| 679J2 | Health education - structured patient focused asthma discuss |
| 663w. | Asthma limits walking up hills or stairs |
| H33z. | Hyperreactive airways disease |
| 9OJ8. | Asthma monitor phone invite |
| 1783 | Asthma trigger - warm air |
| 663P0 | Asthma limits activities 1 to 2 times per month |
| 663N1 | Asthma disturbs sleep weekly |
| H331z | Intrinsic asthma NOS |
| 178A. | Asthma trigger - airborne dust |
| 663U. | Asthma management plan given |
| 9OJA. | Asthma monitored |
| 66YJ. | Asthma annual review |
| 178.. | Asthma trigger |
| 8791 | Further asthma - drug prevent. |
| 663p. | Asthma treatment compliance unsatisfactory |
| 9OJ1. | Attends asthma monitoring |
| 38DT. | Asthma control questionnaire |
| H330. | Extrinsic (atopic) asthma |
| 679J0 | Health education - asthma self management |
| 9OJ.. | Asthma clinic administration |
| 1786 | Asthma trigger - animals |
| 663x. | Asthma limits walking on the flat |
| 388t0 | Royal College Physician asthma assessment 3 question score |
| 66Yp. | Asthma review using Roy Colleg of Physicians three questions |
| 8H2P. | Emergency admission, asthma |
| 663V0 | Occasional asthma |
| 8796 | Asthma control step 3 |
| 663m. | Asthma accident and emergency attendance since last visit |
| 66Ys. | Asthma never causes night symptoms |
| 9OJZ. | Asthma monitoring admin.NOS |
| H3310 | Intrinsic asthma without status asthmaticus |
| 9OJ.. | Asthma monitoring admin. |
| H335. | Chronic asthma with fixed airflow obstruction |
| 38DL. | Asthma control test |
| 663e. | Asthma restricts exercise |
| 9OJ6. | Asthma monitor 3rd letter |
| 663u. | Asthma causes daytime symptoms 1 to 2 times per week |
| 66YP. | Asthma night-time symptoms |
| 1781 | Asthma trigger - pollen |
| 1789 | Asthma trigger - respiratory infection |
| 663N0 | Asthma causing night waking |
| 173A. | Exercise induced asthma |
| 1O2.. | Asthma confirmed |
| 38DV. | Mini asthma quality of life questionnaire |
| 663n. | Asthma treatment compliance satisfactory |
| H33.. | Asthma |
| H3301 | Extrinsic asthma with status asthmaticus |
| 66Yr. | Asthma causes symptoms most nights |
| H33zz | Allergic bronchitis NEC |
| H33zz | Allergic asthma NEC |
| 66Y5. | Change in asthma management plan |
| H33zz | Exercise induced asthma |
| 663f. | Asthma never restricts exercise |
| 38QM. | Childhood Asthma Control Test |
| 663v. | Asthma causes daytime symptoms most days |
| H33z0 | Status asthmaticus NOS |
| 1784 | Asthma trigger - emotion |
| 9OJ7. | Asthma monitor verbal invite |
| H47y0 | Detergent asthma |
| H330z | Extrinsic asthma NOS |
| 663O0 | Asthma never disturbs sleep |
| 66YE. | Asthma monitoring due |
| 66Yu. | Number days absent from school due to asthma in past 6 month |
| 178B. | Asthma trigger - exercise |
| 9OJA. | Asthma monitoring check done |
| H35y7 | Wood asthma |
| 8798 | Asthma control step 5 |
| H330. | Pollen asthma |
| H330. | Hay fever with asthma |
| 66YZ. | Does not have asthma management plan |
| H330. | Childhood asthma |
| H330. | Allergic asthma |
| 663e1 | Asthma severely restricts exercise |
| H333. | Acute exacerbation of asthma |
| 8CR0. | Asthma clinical management plan |
| H33z0 | Severe asthma attack |
| 1787 | Asthma trigger - seasonal |
| 9OJ4. | Asthma monitor 1st letter |
| H3120 | Chronic asthmatic bronchitis |
| 663s. | Asthma never causes daytime symptoms |
| 66YR. | Asthma monitoring by doctor |
| 663P. | Asthma limiting activities |
| 661M1 | Asthma self-management plan agreed |
| 663P2 | Asthma limits activities most days |
| 663V3 | Severe asthma |
| 8CMA0 | Patient has a written asthma personal action plan |

Supplementary table 6: Read codes (v2) used to identify cardiovascular from the primary care data (GP)

| **Read code** | **Description** |
| --- | --- |
| 14A3. | H/O: myocardial infarct <60 |
| 14A4. | H/O: myocardial infarct >60 |
| 14A5. | H/O: angina pectoris |
| 14A6. | H/O: heart failure |
| 14AH. | H/O: Myocardial infarction in last year |
| 14AJ. | H/O: Angina in last year |
| 14AM. | H/O: Heart failure in last year |
| 14AN. | H/O: atrial fibrillation |
| 14AR. | History of atrial flutter |
| 14AT. | History of myocardial infarction |
| 14AW. | H/O acute coronary syndrome |
| 14NB. | H/O: Peripheral vascular disease procedure |
| 1J60. | Suspected heart failure |
| 1O1.. | Heart failure confirmed |
| 21264 | Heart failure resolved |
| 323.. | ECG: myocardial infarction |
| 3232. | ECG: old myocardial infarction |
| 323Z. | ECG: myocardial infarct NOS |
| 3272. | ECG: atrial fibrillation |
| 3273. | ECG: atrial flutter |
| 388D. | New York Heart Assoc classification heart failure symptoms |
| 661M5 | Heart failure self-management plan agreed |
| 662p. | Heart failure 6 month review |
| 662S. | Atrial fibrillation monitoring |
| 662T. | Congestive heart failure monitoring |
| 662W. | Heart failure annual review |
| 679W1 | Education about deteriorating heart failure |
| 679X. | Heart failure education |
| 67D4. | Heart failure information given to patient |
| 68B6. | Heart failure screen |
| 6A9.. | Atrial fibrillation annual review |
| 790D7 | Replacement of valved cardiac conduit |
| 7910. | Plastic repair of mitral valve |
| 79100 | Allograft replacement of mitral valve |
| 79101 | Xenograft replacement of mitral valve |
| 79102 | Prosthetic replacement of mitral valve |
| 79103 | Replacement of mitral valve NEC |
| 79104 | Mitral valvuloplasty NEC |
| 7910y | Other specified plastic repair of mitral valve |
| 7910z | Plastic repair of mitral valve NOS |
| 7911. | Plastic repair of aortic valve |
| 79110 | Allograft replacement of aortic valve |
| 79111 | Xenograft replacement of aortic valve |
| 79112 | Prosthetic replacement of aortic valve |
| 79113 | Replacement of aortic valve NEC |
| 79114 | Aortic valvuloplasty NEC |
| 79115 | Transapical aortic valve implantation |
| 79116 | Transluminal aortic valve implantation |
| 7911y | Other specified plastic repair of aortic valve |
| 7911z | Plastic repair of aortic valve NOS |
| 7914. | Plastic repair of unspecified valve of heart |
| 79140 | Allograft replacement of valve of heart NEC |
| 79141 | Xenograft replacement of valve of heart NEC |
| 79142 | Prosthetic replacement of valve of heart NEC |
| 79143 | Replacement of valve of heart NEC |
| 79146 | Replacement of truncal valve |
| 79150 | Revision of plastic repair of mitral valve |
| 79151 | Revision of plastic repair of aortic valve |
| 79160 | Open mitral valvotomy |
| 79170 | Closed mitral valvotomy |
| 79180 | Annuloplasty of mitral valve |
| 79190 | Percutaneous transluminal mitral valvotomy |
| 792.. | Coronary artery operations |
| 7920. | Saphenous vein graft replacement of coronary artery |
| 7921. | Other autograft replacement of coronary artery |
| 7922. | Allograft replacement of coronary artery |
| 7923. | Prosthetic replacement of coronary artery |
| 7924. | Revision of bypass for coronary artery |
| 79240 | Revision of bypass for one coronary artery |
| 79241 | Revision of bypass for two coronary arteries |
| 79242 | Revision of bypass for three coronary arteries |
| 79243 | Revision of bypass for four or more coronary arteries |
| 7924y | Other specified revision of bypass for coronary artery |
| 7924z | Revision of bypass for coronary artery NOS |
| 7925. | Connection of mammary artery to coronary artery |
| 79275 | Open angioplasty of coronary artery |
| 7928. | Transluminal balloon angioplasty of coronary artery |
| 79280 | Percut transluminal balloon angioplasty one coronary artery |
| 79281 | Percut translum balloon angioplasty mult coronary arteries |
| 79282 | Percut translum balloon angioplasty bypass graft coronary a |
| 79283 | Percut translum cutting balloon angioplasty coronary artery |
| 7928y | Transluminal balloon angioplasty of coronary artery OS |
| 7928z | Transluminal balloon angioplasty of coronary artery NOS |
| 79290 | Percutaneous transluminal laser coronary angioplasty |
| 79293 | Rotary blade coronary angioplasty |
| 79294 | Insertion of coronary artery stent |
| 79295 | Insertion of drug-eluting coronary artery stent |
| 792D. | Other bypass of coronary artery |
| 792Dy | Other specified other bypass of coronary artery |
| 792Dz | Other bypass of coronary artery NOS |
| 7936A | Implant intravenous pacemaker for atrial fibrillation |
| 793G. | Perc translumin balloon angioplasty stenting coronary artery |
| 793Gy | OS perc translumina balloon angioplast stenting coronary art |
| 793Gz | Perc translum balloon angioplasty stenting coronary art NOS |
| 793M1 | Perc transluminal ablation of atrial wall for atrial flutter |
| 793M2 | Percutaneous transluminal internal cardioversion NEC |
| 793M3 | Perc translum ablat conduct sys heart for atrial flutter NEC |
| 7L1H. | External resuscitation |
| 7L1H0 | Direct current cardioversion |
| 7L1H1 | External cardioversion NEC |
| 7L1H2 | Internal electrode cardioversion |
| 7L1H8 | Chemical cardioversion |
| 840.. | Direct current cardioversion planned |
| 889A. | Diab mellit insulin-glucose infus acute myocardial infarct |
| 8CeC. | Preferred place of care for next exacerbation heart failure |
| 8CL3. | Heart failure care plan discussed with patient |
| 8CMK. | Has heart failure management plan |
| 8CMW2 | Atrial fibrillation care pathway |
| 8CMW8 | Heart failure clinical pathway |
| 8H2S. | Admit heart failure emergency |
| 8H44. | Cardiological referral |
| 8H440 | Referral to cardiology multidisciplinary team |
| 8H5G. | Referral to Cardiothoracic surgeon |
| 8H7v. | Referral to cardiac rehabilitation nurse |
| 8HBE. | Heart failure follow-up |
| 8HBJ. | Stroke / transient ischaemic attack referral |
| 8Hg8. | Discharge from practice nurse heart failure clinic |
| 8HgD. | Discharge from heart failure nurse service |
| 8HHb. | Referral to heart failure nurse |
| 8HHM. | Ref to multidisciplinary stroke function improvement service |
| 8HHW. | Referral for warfarin monitoring |
| 8HHz. | Referral to heart failure exercise programme |
| 8Hk0. | Referred to heart failure education group |
| 8Hkk. | Referral to cardiac rehabilitation programme |
| 8Hkl. | Referral to cardiac rehabilitation service by secondary care |
| 8Hkt. | Referral to community cardiology service |
| 8HQ7. | Referral for echocardiography |
| 8HR9. | Referral for 24 hour ECG |
| 8HRA. | Referral for exercise ECG |
| 8HRD. | Referral for ambulatory electrocardiogram |
| 8HRF. | Referral for cardiac pacemaker check |
| 8HRG. | Referral for cardiac event recording |
| 8HTL. | Referral to heart failure clinic |
| 8HTL0 | Referral to rapid access heart failure clinic |
| 8HTQ. | Referral to stroke clinic |
| 8HTs. | Referral to community anticoagulation clinic |
| 8HTy. | Referral to atrial fibrillation clinic |
| 8HVE. | Private referral cardiothoracic surgeon |
| 8HVJ. | Private referral to cardiologist |
| 8IB8. | Referral to heart failure exercise programme not indicated |
| 8IE1. | Referral to heart failure exercise programme declined |
| 8L40. | Coronary artery bypass graft operation planned |
| 8L41. | Coronary angioplasty planned |
| 8OAD. | Provision of written information about atrial fibrillation |
| 9hF.. | Exception reporting: atrial fibrillation quality indicators |
| 9hF1. | Excepted from atrial fibrillation qual indic: Inform dissent |
| 9hH.. | Exception reporting: heart failure quality indicators |
| 9hH0. | Excepted heart failure quality indicators: Patient unsuitabl |
| 9hH1. | Excepted heart failure quality indicators: Informed dissent |
| 9hS.. | Exception report: peripherl arterial disease quality indicat |
| 9m5.. | High risk of heart failure screening invitation |
| 9N0k. | Seen in heart failure clinic |
| 9N2p. | Seen by community heart failure nurse |
| 9N4s. | Did not attend practice nurse heart failure clinic |
| 9N4w. | Did not attend heart failure clinic |
| 9N6T. | Referred by heart failure nurse specialist |
| 9Or.. | Heart failure monitoring administration |
| 9Or0. | Heart failure review completed |
| 9Or1. | Heart failure monitoring telephone invite |
| 9Or2. | Heart failure monitoring verbal invite |
| 9Or3. | Heart failure monitoring first letter |
| 9Or4. | Heart failure monitoring second letter |
| 9Or5. | Heart failure monitoring third letter |
| 9Os.. | Atrial fibrillation monitoring administration |
| 9Os0. | Atrial fibrillation monitoring first letter |
| 9Os1. | Atrial fibrillation monitoring second letter |
| 9Os2. | Atrial fibrillation monitoring third letter |
| 9Os3. | Atrial fibrillation monitoring verbal invite |
| 9Os4. | Atrial fibrillation monitoring telephone invite |
| G11.. | Mitral valve diseases |
| G110. | Mitral stenosis |
| G112. | Mitral stenosis with insufficiency |
| G113. | Nonrheumatic mitral valve stenosis |
| G114. | Ruptured mitral valve cusp |
| G11z. | Mitral valve disease NOS |
| G12z. | Rheumatic aortic valve disease NOS |
| G13.. | Diseases of mitral and aortic valves |
| G130. | Mitral and aortic stenosis |
| G131. | Mitral stenosis and aortic insufficiency |
| G132. | Mitral insufficiency and aortic stenosis |
| G133. | Mitral and aortic incompetence |
| G13y. | Multiple mitral and aortic valve involvement |
| G13z. | Mitral and aortic valve disease NOS |
| G1yz1 | Rheumatic left ventricular failure |
| G232. | Hypertensive heart&renal dis wth (congestive) heart failure |
| G3... | Ischaemic heart disease |
| G30.. | Acute myocardial infarction |
| G301. | Other specified anterior myocardial infarction |
| G301z | Anterior myocardial infarction NOS |
| G304. | Posterior myocardial infarction NOS |
| G305. | Lateral myocardial infarction NOS |
| G306. | True posterior myocardial infarction |
| G3071 | Acute non-ST segment elevation myocardial infarction |
| G308. | Inferior myocardial infarction NOS |
| G30B. | Acute posterolateral myocardial infarction |
| G30X. | Acute transmural myocardial infarction of unspecif site |
| G30X0 | Acute ST segment elevation myocardial infarction |
| G30y. | Other acute myocardial infarction |
| G30yz | Other acute myocardial infarction NOS |
| G30z. | Acute myocardial infarction NOS |
| G31.. | Other acute and subacute ischaemic heart disease |
| G310. | Postmyocardial infarction syndrome |
| G311. | Preinfarction syndrome |
| G3110 | Myocardial infarction aborted |
| G3111 | Unstable angina |
| G3113 | Refractory angina |
| G3115 | Acute coronary syndrome |
| G31y. | Other acute and subacute ischaemic heart disease |
| G31y0 | Acute coronary insufficiency |
| G31yz | Other acute and subacute ischaemic heart disease NOS |
| G32.. | Old myocardial infarction |
| G33.. | Angina pectoris |
| G331. | Prinzmetal's angina |
| G33z. | Angina pectoris NOS |
| G33z3 | Angina on effort |
| G33z5 | Post infarct angina |
| G33z7 | Stable angina |
| G33zz | Angina pectoris NOS |
| G34.. | Other chronic ischaemic heart disease |
| G340. | Coronary atherosclerosis |
| G34y. | Other specified chronic ischaemic heart disease |
| G34yz | Other specified chronic ischaemic heart disease NOS |
| G34z. | Other chronic ischaemic heart disease NOS |
| G35.. | Subsequent myocardial infarction |
| G350. | Subsequent myocardial infarction of anterior wall |
| G351. | Subsequent myocardial infarction of inferior wall |
| G353. | Subsequent myocardial infarction of other sites |
| G35X. | Subsequent myocardial infarction of unspecified site |
| G36.. | Certain current complication follow acute myocardial infarct |
| G38.. | Postoperative myocardial infarction |
| G380. | Postoperative transmural myocardial infarction anterior wall |
| G381. | Postoperative transmural myocardial infarction inferior wall |
| G383. | Postoperative transmural myocardial infarction unspec site |
| G384. | Postoperative subendocardial myocardial infarction |
| G38z. | Postoperative myocardial infarction, unspecified |
| G3y.. | Other specified ischaemic heart disease |
| G3z.. | Ischaemic heart disease NOS |
| G540. | Mitral valve incompetence |
| G5400 | Mitral incompetence, non-rheumatic |
| G5401 | Mitral incompetence, cause unspecified |
| G5402 | Mitral valve prolapse |
| G5403 | Mitral valve leaf prolapse |
| G540z | Mitral valve disorders NOS |
| G541. | Aortic valve disorders |
| G5414 | Aortic valve stenosis with insufficiency |
| G541z | Aortic valve disorders NOS |
| G544. | Multiple valve diseases |
| G5441 | Disorders of both mitral and tricuspid valves |
| G5442 | Combined disorders of mitral, aortic and tricuspid valves |
| G544X | Multiple valve disease, unspecified |
| G573. | Atrial fibrillation and flutter |
| G5730 | Atrial fibrillation |
| G5731 | Atrial flutter |
| G5732 | Paroxysmal atrial fibrillation |
| G5733 | Non-rheumatic atrial fibrillation |
| G5734 | Permanent atrial fibrillation |
| G5735 | Persistent atrial fibrillation |
| G5736 | Paroxysmal atrial flutter |
| G573z | Atrial fibrillation and flutter NOS |
| G58.. | Heart failure |
| G580. | Congestive heart failure |
| G5800 | Acute congestive heart failure |
| G5801 | Chronic congestive heart failure |
| G5804 | Congestive heart failure due to valvular disease |
| G581. | Left ventricular failure |
| G5810 | Acute left ventricular failure |
| G582. | Acute heart failure |
| G583. | Heart failure with normal ejection fraction |
| G584. | Right ventricular failure |
| G58z. | Heart failure NOS |
| G5y4z | Post cardiac operation heart failure NOS |
| G670. | Cerebral atherosclerosis |
| G677. | Occlusion/stenosis cerebral arts not result cerebral infarct |
| G7... | Arterial, arteriole and capillary disease |
| G70.. | Atherosclerosis |
| G700. | Aortic atherosclerosis |
| G701. | Renal artery atherosclerosis |
| G702. | Extremity artery atheroma |
| G702z | Extremity artery atheroma NOS |
| G70y. | Other specified artery atheroma |
| G70y0 | Carotid artery atherosclerosis |
| G73.. | Other peripheral vascular disease |
| G734. | Peripheral arterial disease |
| G73y. | Other specified peripheral vascular disease |
| G73yz | Other specified peripheral vascular disease NOS |
| G73z. | Peripheral vascular disease NOS |
| G73z0 | Intermittent claudication |
| G73z1 | Spasm of peripheral artery |
| G73zz | Peripheral vascular disease NOS |
| G7y.. | Other specified arterial, arteriole or capillary disease |
| G7z.. | Arterial, arteriole and capillary diseases NOS |
| Gyu10 | [X]Other mitral valve diseases |
| Gyu3. | [X]Ischaemic heart diseases |
| Gyu30 | [X]Other forms of angina pectoris |
| Gyu32 | [X]Other forms of acute ischaemic heart disease |
| Gyu33 | [X]Other forms of chronic ischaemic heart disease |
| Gyu34 | [X]Acute transmural myocardial infarction of unspecif site |
| Gyu36 | [X]Subsequent myocardial infarction of unspecified site |
| Gyu55 | [X]Other nonrheumatic mitral valve disorders |
| Gyu56 | [X]Other aortic valve disorders |
| Gyu5A | [X]Aortic valve disorders in diseases classified elsewhere |
| Gyu5D | [X]Multiple valve disorders/diseases CE |
| Gyu70 | [X]Atherosclerosis of other arteries |
| Gyu74 | [X]Other specified peripheral vascular diseases |
| P641. | Bicuspid aortic valve |
| P65.. | Congenital mitral stenosis |
| P650. | Congenital mitral stenosis, unspecified |
| P652. | Parachute deformity of the mitral valve |
| P65z. | Congenital mitral stenosis NOS |
| P66.. | Congenital mitral insufficiency |
| SP002 | Mechanical complication of heart valve prosthesis |
| SP003 | Mechanical complication of coronary bypass |
| SP076 | Coronary artery bypass graft occlusion |
| SP084 | Heart transplant failure and rejection |
| SP085 | Heart-lung transplant failure and rejection |
| SP111 | Cardiac insufficiency as a complication of care |
| SyuK6 | [X]Oth complics of cardiac & vasc prosth devices/impl/graft |
| TB012 | Implant of heart valve prosthesis + complication, no blame |
| ZV433 | [V]Has artificial heart valve |
| ZV457 | [V]Presence of aortocoronary bypass graft |
| ZV458 | [V]Presence of coronary angioplasty implant and graft |
| ZV45H | [V]Presence of prosthetic heart valve |
| ZV45K | [V]Presence of coronary artery bypass graft |
| ZV45L | [V]Status following coronary angioplasty NOS |
| ZVu6e | [X]Presence of other heart valve replacement |

Supplementary table 7: ICD-10 version:2019 codes used to identify depression from the hospital admissions data (PEDW)

| **ICD-10 code** | **Description** |
| --- | --- |
| F32 | Depressive episode |
| F32.0 | Mild depressive episode |
| F32.1 | Moderate depressive episode |
| F32.2 | Sever depressive episode without psychotic symptoms |
| F32.3 | Sever depressive episode with psychotic symptoms |
| F32.8 | Other depressive episodes |
| F32.9 | Depressive episode, unspecified |
| F33 | Recurrent depressive disorder |
| F33.0 | Recurrent depressive disorder, current episode mild |
| F33.1 | Recurrent depressive disorder, current episode moderate |
| F33.2 | Recurrent depressive disorder, current episode sever without psychotic symptoms |
| F33.3 | Recurrent depressive disorder, current episode sever with psychotic symptoms |
| F33.4 | Recurrent depressive disorder, currently in remission |
| F33.8 | Other recurrent depressive disorders |
| F33.9 | Recurrent depressive episode, unspecified |

Supplementary table 8: ICD-10 version:2019 codes used to identify diabetes from the hospital admissions data (PEDW)

| **ICD-10 code** | **Description** |
| --- | --- |
| E10 | Insulin-dependent diabetes mellitus |
| E100 | Insulin-dependent diabetes mellitus |
| E101 | Insulin-dependent diabetes mellitus |
| E102 | Insulin-dependent diabetes mellitus |
| E103 | Insulin-dependent diabetes mellitus |
| E104 | Insulin-dependent diabetes mellitus |
| E105 | Insulin-dependent diabetes mellitus |
| E106 | Insulin-dependent diabetes mellitus |
| E107 | Insulin-dependent diabetes mellitus |
| E108 | Insulin-dependent diabetes mellitus |
| E109 | Insulin-dependent diabetes mellitus |
| O240 | Diabetes mellitus in pregnancy: Pre-existing diabetes mellitus, insulin-dependent |
| E11 | Non-insulin-dependent diabetes mellitus |
| E110 | Non-insulin-dependent diabetes mellitus |
| E111 | Non-insulin-dependent diabetes mellitus |
| E112 | Non-insulin-dependent diabetes mellitus |
| E113 | Non-insulin-dependent diabetes mellitus |
| E114 | Non-insulin-dependent diabetes mellitus |
| E115 | Non-insulin-dependent diabetes mellitus |
| E116 | Non-insulin-dependent diabetes mellitus |
| E117 | Non-insulin-dependent diabetes mellitus |
| E118 | Non-insulin-dependent diabetes mellitus |
| E119 | Non-insulin-dependent diabetes mellitus |
| O241 | Diabetes mellitus in pregnancy: Pre-existing diabetes mellitus, non-insulin-dependent |
| E13 | Other specified diabetes mellitus |
| E130 | Other specified diabetes mellitus |
| E131 | Other specified diabetes mellitus |
| E132 | Other specified diabetes mellitus |
| E133 | Other specified diabetes mellitus |
| E134 | Other specified diabetes mellitus |
| E135 | Other specified diabetes mellitus |
| E136 | Other specified diabetes mellitus |
| E137 | Other specified diabetes mellitus |
| E138 | Other specified diabetes mellitus |
| E139 | Other specified diabetes mellitus |
| E14 | Unspecified diabetes mellitus |
| E140 | Unspecified diabetes mellitus |
| E141 | Unspecified diabetes mellitus |
| E142 | Unspecified diabetes mellitus |
| E143 | Unspecified diabetes mellitus |
| E144 | Unspecified diabetes mellitus |
| E145 | Unspecified diabetes mellitus |
| E146 | Unspecified diabetes mellitus |
| E147 | Unspecified diabetes mellitus |
| E148 | Unspecified diabetes mellitus |
| E149 | Unspecified diabetes mellitus |
| O243 | Diabetes mellitus in pregnancy: Pre-existing diabetes mellitus, unspecified |
| O249 | Diabetes mellitus in pregnancy, unspecified |
| E12 | Malnutrition-related diabetes mellitus |
| E120 | Malnutrition-related diabetes mellitus |
| E121 | Malnutrition-related diabetes mellitus |
| E122 | Malnutrition-related diabetes mellitus |
| E123 | Malnutrition-related diabetes mellitus |
| E124 | Malnutrition-related diabetes mellitus |
| E125 | Malnutrition-related diabetes mellitus |
| E126 | Malnutrition-related diabetes mellitus |
| E127 | Malnutrition-related diabetes mellitus |
| E128 | Malnutrition-related diabetes mellitus |
| E129 | Malnutrition-related diabetes mellitus |
| O242 | Diabetes mellitus in pregnancy: Pre-existing malnutrition-related diabetes mellitus |

Supplementary table 9: ICD-10 version:2019 codes used to identify asthma from the hospital admissions data (PEDW)

| **ICD-10 code** | **Description** |
| --- | --- |
| J45 | Asthma |
| J450 | Predominantly allergic asthma |
| J451 | Nonallergic asthma |
| J458 | Mixed asthma |
| J459 | Asthma, unspecified |
| J46X | Status asthmaticus |

Supplementary table 10: ICD-10 version:2019 codes used to identify cardiovascular from the hospital admissions data (PEDW)

| **ICD-10 code** | **Description** |
| --- | --- |
| I21 | Acute myocardial infarction |
| I21.0 | Acute transmural myocardial infarction of anterior wall |
| I21.1 | Acute transmural myocardial infarction of inferior wall |
| I21.2 | Acute transmural myocardial infarction of other sites |
| I21.3 | Acute transmural myocardial infarction of unspecified site |
| I21.4 | Acute subendocardial myocardial infarction |
| I21.9 | Acute myocardial infarction, unspecified |
| I22 | Subsequent myocardial infarction |
| I22.0 | Subsequent myocardial infarction of anterior wall |
| I22.1 | Subsequent myocardial infarction of inferior wall |
| I22.8 | Subsequent myocardial infarction of other sites |
| I22.9 | Subsequent myocardial infarction of unspecified site |
| I23 | Certain current complications following acute myocardial infarction |
| I23.0 | Haemopericardium as current complication following acute myocardial infarction |
| I23.1 | Atrial septal defect as current complication following acute myocardial infarction |
| I23.2 | Ventricular septal defect as current complication following acute myocardial infarction |
| I23.3 | Rupture of cardiac wall without haemopericardium as current complication following acute myocardial |
| I23.4 | Rupture of chordae tendineae as current complication following acute myocardial infarction |
| I23.5 | Rupture of papillary muscle as current complication following acute myocardial infarction |
| I23.6 | Thrombosis of atrium, auricular appendage, and ventricle as current complications following acute my |
| I23.8 | Other current complications following acute myocardial infarction |
| I63 | Cerebral infarction |
| I63.0 | Cerebral infarction due to thrombosis of precerebral arteries |
| I63.1 | Cerebral infarction due to embolism of precerebral arteries |
| I63.2 | Cerebral infarction due to unspecified occlusion or stenosis of precerebral arteries |
| I63.3 | Cerebral infarction due to thrombosis of cerebral arteries |
| I63.4 | Cerebral infarction due to embolism of cerebral arteries |
| I63.5 | Cerebral infarction due to unspecified occlusion or stenosis of cerebral arteries |
| I63.6 | Cerebral infarction due to cerebral venous thrombosis, nonpyogenic |
| I63.8 | Other cerebral infarction |
| I63.9 | Cerebral infarction, unspecified |
| I64 | Stroke, not specified as haemorrhage or infarction |
| I64.0 | Stroke, not specified as haemorrhage or infarction |

Supplementary table 11. Estimates of time to vaccination by depression, diabetes, asthma, and cardiovascular independently

| Condition | | Median Time | | | P value |
| --- | --- | --- | --- | --- | --- |
|  |  | **Estimate** | **Std. Error** | **95% CI** |  |
| Depression | 0 | 125 | 1.36 | 122.33 – 127.67 | 0.127 |
|  | 1 | 123 | 2.55 | 118.00 – 127.99 |  |
| Diabetes | 0 | 125 | 1.29 | 122.47 – 127.53 | 0.341 |
|  | 1 | 120 | 4.71 | 110.76 – 129.24 |  |
| Asthma | 0 | 126 | 1.44 | 123.17 – 128.83 | 0.323 |
|  | 1 | 119 | 2.38 | 114.34 – 123.66 |  |
| Cardiovascular | 0 | 125 | 1.28 | 122.50 – 127.51 | 0.402 |
|  | 1 | 125 | 5.78 | 113.66 – 136.34 |  |

Supplementary table 12. Cox Regression analysis of depression, demographics, and smoking status factors among pregnant women eligible for vaccination, adjusted analysis.

| **Characteristic** | | **HR^1^ (95% CI^2^)** | ***P* value^3^** |
| --- | --- | --- | --- |
| **Age** | 25-29 | Reference |  |
|  | 18-24 | .99 (.92 – 1.06) | .752 |
|  | 30-39 | 1.17 (1.12 – 1.24) | <.001 |
|  | 40-50 | 1.33 (1.18 – 1.49) | <.001 |
| **Ethnic groups** | White | Reference |  |
|  | Asian | 1.09 (.97–1.22) | .131 |
|  | Other | 1.16 (1.01–1.34) | .041 |
|  | Mixed | 1.02 (.81–1.28) | .891 |
|  | Black | 1.06 (.87–1.29) | .549 |
|  | Unknown | .94 (.87–1.01) | .072 |
| **WIMD quintile 2019** | 5^th^ (Least deprived) | Reference |  |
|  | 4^th^ | .90 (.84 – .97) | .005 |
|  | 3^rd^ | .82 (.76 – .88) | <.001 |
|  | 2^nd^ | .92 (.85 – .98) | .016 |
|  | 1^st^ (Most deprived) | 089 (.82 – .96) | .002 |
| **Smoking status** | Never Smoker | Reference |  |
|  | Former Smoker | .91 (.85 – .98) | .013 |
|  | Current Smoker | .87 (.81 – .93) | <.001 |
|  | Unknown | 1.09 (1.03 – 1.15) | .002 |
| **Depression** | No | Reference |  |
|  | Yes | 1.08 (1.03 – 1.14) | .002 |

^1^Hazard Ratio, ^2^Confidence Interval (95%), ^3^significance level accepted at <0.05

Supplementary table 13. Cox Regression analysis of diabetes, demographics, and smoking status factors among pregnant women eligible for vaccination, adjusted analysis.

| **Characteristic** | | **HR^1^ (95% CI^2^)** | ***P* value^3^** |
| --- | --- | --- | --- |
| **Age** | 25-29 | Reference |  |
|  | 18-24 | .99 (.92 – 1.07) | .793 |
|  | 30-39 | 1.17 (1.12 – 1.24) | <.001 |
|  | 40-50 | 1.33 (1.19 – 1.49) | <.001 |
| **Ethnic groups** | White | Reference |  |
|  | Asian | 1.08 (.96–1.21) | .189 |
|  | Other | 1.15 (.99–1.32) | .063 |
|  | Mixed | 1.01 (.80–1.27) | .927 |
|  | Black | 1.05 (.86–1.28) | .636 |
|  | Unknown | .93 (.87–1.00) | .049 |
| **WIMD quintile 2019** | 5^th^ (Least deprived) | Reference |  |
|  | 4^th^ | .90 (.84 – .97) | .005 |
|  | 3^rd^ | .82 (.76 – .88) | <.001 |
|  | 2^nd^ | .92 (.86 – .97) | .023 |
|  | 1^st^ (Most deprived) | 0.90 (.83 – .97) | .004 |
| **Smoking status** | Never Smoker | Reference |  |
|  | Former Smoker | .92 (.86 – .99) | .024 |
|  | Current Smoker | .88 (.82 – .95) | <.001 |
|  | Unknown | 1.09 (1.03 – 1.15) | .003 |
| **Diabetes** | No | Reference |  |
|  | Yes | 1.01 (.93 – 1.11) | .761 |

Supplementary table 14. Cox Regression analysis of asthma, demographics, and smoking status factors among pregnant women eligible for vaccination, adjusted analysis.

| **Characteristic** | | **HR^1^ (95% CI^2^)** | ***P* value^3^** |
| --- | --- | --- | --- |
| **Age** | 25-29 | Reference |  |
|  | 18-24 | .99 (.92 – 1.07) | .804 |
|  | 30-39 | 1.18 (1.12 – 1.24) | <.001 |
|  | 40-50 | 1.33 (1.19 – 1.50) | <.001 |
| **Ethnic groups** | White | Reference |  |
|  | Asian | 1.09 (.97–1.22) | .145 |
|  | Other | 1.16 (1.00–1.34) | .049 |
|  | Mixed | 1.02 (.81–1.28) | .888 |
|  | Black | 1.05 (.87–1.28) | .596 |
|  | Unknown | .93 (.87–1.00) | .054 |
| **WIMD quintile 2019** | 5^th^ (Least deprived) | Reference |  |
|  | 4^th^ | .90 (.84 – .97) | .005 |
|  | 3^rd^ | .82 (.76 – .88) | <.001 |
|  | 2^nd^ | .92 (.85 – .99) | .021 |
|  | 1^st^ (Most deprived) | 0.90 (.83 – .96) | .003 |
| **Smoking status** | Never Smoker | Reference |  |
|  | Former Smoker | .92 (.86 – .99) | .022 |
|  | Current Smoker | .88 (.82 – .94) | <.001 |
|  | Unknown | 1.09 (1.03 – 1.15) | .002 |
| **Asthma** | No | Reference |  |
|  | Yes | 1.05 (0.99– 1.10) | .073 |

Supplementary table 15. Cox Regression analysis of cardiovascular, demographics, and smoking status factors among pregnant women eligible for vaccination, adjusted analysis.

| **Characteristic** | | **HR^1^ (95% CI^2^)** | ***P* value^3^** |
| --- | --- | --- | --- |
| **Age** | 25-29 | Reference |  |
|  | 18-24 | .99 (.92 – 1.07) | .795 |
|  | 30-39 | 1.17 (1.12 – 1.24) | <.001 |
|  | 40-50 | 1.33 (1.19 – 1.50) | <.001 |
| **Ethnic groups** | White | Reference |  |
|  | Asian | 1.08 (.97–1.21) | .175 |
|  | Other | 1.15 (.99–1.33) | .059 |
|  | Mixed | 1.01 (.81–1.27) | .916 |
|  | Black | 1.05 (.86–1.28) | .628 |
|  | Unknown | .93 (.87–1.00) | .050 |
| **WIMD quintile 2019** | 5^th^ (Least deprived) | Reference |  |
|  | 4^th^ | .90 (.84 – .97) | .004 |
|  | 3^rd^ | .82 (.76 – .88) | <.001 |
|  | 2^nd^ | .92 (.86 – .99) | .023 |
|  | 1^st^ (Most deprived) | 0.90 (.83 – .97) | .004 |
| **Smoking status** | Never Smoker | Reference |  |
|  | Former Smoker | .92 (.86 – .99) | .024 |
|  | Current Smoker | .88 (.82 – .95) | <.001 |
|  | Unknown | 1.09 (1.03 – 1.15) | .002 |
| **Cardiovascular** | No | Reference |  |
|  | Yes | 1.06 (0.94– 1.19) | .367 |

Supplementary table 16. Bootstrapping internal validation of depression and other factors associated with vaccination uptake among pregnant women eligible for vaccination

| **Characteristic** | | **B^1^ (BCa 95% CI^2^)** | **Bias** | **Std. Error^3^** | ***P* value^4^** |
| --- | --- | --- | --- | --- | --- |
| **Age** | 18-24 | -.01 (-.09 – .06) | .000 | .038 | .784 |
|  | 30-39 | .16 (.11 – .21) | .000 | .026 | <.001 |
|  | 40-50 | .28 (.16 – .40) | .000 | .058 | <.001 |
| **Ethnic groups** | Asian | .09 (-.03 – .21) | .000 | .059 | .137 |
|  | Other | .15 (-.01 – .29) | .004 | .073 | .037 |
|  | Mixed | .02 (-.21 – .25) | .000 | .120 | .890 |
|  | Black | .06 (-.12 – .25) | -.001 | .098 | .050 |
|  | Unknown | -.07 (-.13 – .001) | .-.001 | .035 | .063 |
| **WIMD quintile 2019** | 4^th^ | -.11 (-.18 – -.03) | -.001 | .037 | .003 |
|  | 3^rd^ | -.20 (-.28 – -.13) | .000 | .038 | <.001 |
|  | 2^nd^ | -.09 (-.16 – -.02) | .002 | .036 | .013 |
|  | 1^st^ (Most deprived) | -.12 (-.19 – -.04) | .002 | .038 | .<.001 |
| **Smoking status** | Former Smoker | -.09 (-.17 - -.02) | .000 | .037 | .015 |
|  | Current Smoker | -.14 (-.22 - -.07) | -.001 | .039 | <.001 |
|  | Unknown | .09 (.03 - .14) | .000 | .027 | .002 |
| **Depression** | Multimorbidity | .08 (.03 - .13) | .001 | .027 | .004 |

^1^Bootstrapped Beta Coefficient, ^2^BCa Confidence Interval (95%), ^3^Standard Error, ^4^significance level accepted at <0.05
